# MS4A4A modifies the risk of Alzheimer disease by regulating lipid metabolism and immune response in a unique microglia state

**DOI:** 10.1101/2023.02.06.23285545

**Authors:** Shih-Feng You, Logan Brase, Fabia Filipello, Abhirami K. Iyer, Jorge Del-Aguila, June He, Ricardo D’Oliveira Albanus, John Budde, Joanne Norton, Jen Gentsch, Nina M. Dräger, Sydney M. Sattler, Martin Kampmann, Laura Piccio, John C. Morris, Richard J. Perrin, Eric McDade, Dominantly Inherited Alzheimer Network, Steven M. Paul, Anil G. Cashikar, Bruno A. Benitez, Oscar Harari, Celeste M. Karch

**Affiliations:** Department of Psychiatry, Washington University in St. Louis School of Medicine, USA; Institute for Neurodegenerative Diseases, Department of Biochemistry and Biophysics, University of California, San Francisco, San Francisco, CA, USA; Charles Perkins Centre and Brain and Mind Centre, School of Medical Sciences, University of Sydney, Sydney, NSW, Australia; Department of Neurology, Washington University in St. Louis School of Medicine, USA; The Charles F. and Joanne Knight Alzheimer Disease Research Center, Washington University School of Medicine, St. Louis, Missouri, USA; Department of Pathology and Immunology, Washington University School of Medicine, St. Louis, Missouri, USA; Department of Neurology, Harvard Medical School and Beth Israel Deaconess Medical Center, Boston, Massachusetts, USA; NeuroGenomics and Informatics Center, Washington University School of Medicine, St. Louis, Missouri, USA

## Abstract

Genome-wide association studies (GWAS) have identified many modifiers of Alzheimer disease (AD) risk enriched in microglia. Two of these modifiers are common variants in the *MS4A* locus (rs1582763: protective and rs6591561: risk) and serve as major regulators of CSF sTREM2 levels. To understand their functional impact on AD, we used single nucleus transcriptomics to profile brains from carriers of these variants. We discovered a “chemokine” microglial subpopulation that is altered in *MS4A* variant carriers and for which *MS4A4A* is the major regulator. The protective variant increases *MS4A4A* expression and shifts the chemokine microglia subpopulation to an interferon state, while the risk variant suppresses *MS4A4A* expression and reduces this subpopulation of microglia. Our findings provide a mechanistic explanation for the AD variants in the *MS4A* locus. Further, they pave the way for future mechanistic studies of AD variants and potential therapeutic strategies for enhancing microglia resilience in AD pathogenesis.

## Main

Alzheimer disease (AD) is driven by complex genetic architecture that can be leveraged to understand key pathways that may promote or suppress disease processes. Genome-wide association studies (GWAS) have revealed more than 70 loci associated with AD risk, many of which harbor genes that are enriched in microglia^1-5^. Microglia, the innate immune cells of the brain, produce inflammatory signals in response to environmental stressors. Dynamic changes in microglia function have been reported during AD progression^6^; thus, understanding the factors that influence these states is critical to therapeutically harnessing microglia function.

The microglia-enriched *MS4A* gene family encodes a class of tetraspanin proteins that have been implicated in AD^1,2,7^. Common variants in the *MS4A* locus are associated with AD susceptibility and regulation of soluble TREM2 (sTREM2) levels in cerebrospinal fluid (CSF)^1,2,8-10^. Two independent signals in the *MS4A* locus are associated with altered CSF sTREM2 levels^8-10^. The most significant signal occurs in an intergenic region near *MS4A4A* (rs1582763), which is associated with elevated CSF sTREM2 levels as well as reduced AD risk^2,11^ and delayed age-at-onset (e.g., protective)^12^. A second, independent signal occurs in the coding region of *MS4A4A* (rs6591561; p.M159V) and is associated with lower CSF sTREM2 levels as well as increased AD risk ^2,11^ and accelerated age-at-onset^12^. However, the impact of these AD-modifying variants on microglia function and AD pathophysiology is unknown.

To date, with a few exceptions^5,13-16^, the functional consequences of variants identified in AD risk loci are unknown. We now build upon emerging single nucleus RNA sequencing (snRNA-seq) technologies to look beyond disease status and cell-type proportion to establish a blueprint to pinpoint the function of common variants that drive neurodegenerative disease.

Here, we combined genomic and snRNA-seq technologies to identify the function of AD variants in the *MS4A* gene locus. We discovered that variants in the *MS4A* locus regulate a subpopulation of microglia that are enriched in chemokine function. The AD protective variant drives elevated *MS4A4A* expression and shifts the chemokine microglia subpopulation towards an interferon state, while the AD risk variant suppresses *MS4A4A* expression and the chemokine microglia subpopulation. Together, variants in the *MS4A* locus illustrate the power of microglia to promote or suppress AD pathophysiology.

## Results

### MS4A4A protective variant rs1582763 reduces the inflammatory response and enhances lipid metabolism pathways

To understand the impact of the AD protective variant in the *MS4A* locus (rs1582763), we evaluated gene expression in rs1582763 variant carrier brains (GG, GA, AA; Figure 1A-B). Meta-analysis of trans-eQTLs was performed using three independent AD and control brain cohorts (n=579 brains from the Knight ADRC, Mayo RNAseq Study (Mayo), and the Religious Orders Study and Memory and Aging Project (ROSMAP); Figure 1B and 2A; Supplemental Table 1). After correcting for age, sex, and AD status, we identified 1,071 differentially expressed protein-coding genes (FDR<0.05; Figure 2A; Supplemental Table 2). These genes were enriched in pathways associated with reduced pro-inflammatory responses (NF-kB [p = 6.29×10^−11^], Cytokine [p = 1.12×10^−10^], IFN-γ [p = 4.59×10^−10^], Complement [p = 8.21×10^−8^]); increased anti-inflammatory response (Type I-IFN [p = 1.7×10^−4^]); elevated lipid metabolism (cholesterol efflux [p = 6.54×10^−5^]); and metabolism (mTORC1 [p = 4.05×10^−3^]) (Figure 2B; Supplemental Table 3). Several AD risk genes were significantly increased in the presence of the minor allele of rs1582763: *PILRA, MS4A14, STAG3, C7orf61, EFHD1, CCP110, ATXN3, ESCO2* (Supplemental Table 4).

**Figure 1.**
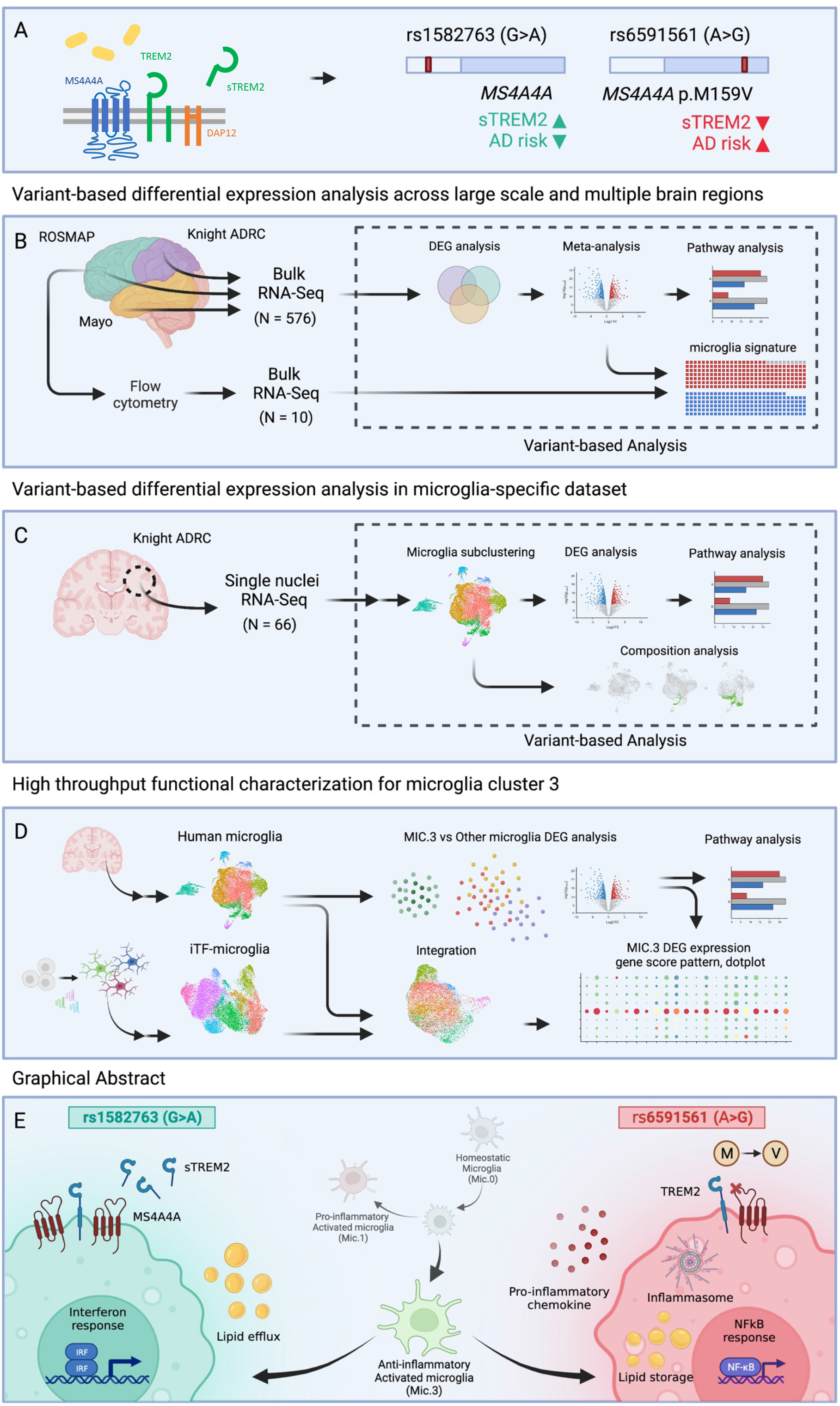
Study overview. A. MS4A4A and TREM2 reside on the plasma membrane in microglia. Prior work demonstrates that rs1582763 is associated with elevated CSF sTREM2 and reduced AD risk (termed: protective) and rs6591561 is associated with reduced CSF sTREM2 and increased AD risk (termed: risk) B. Trans-eQTL analyses for rs1582763 and rs6591561 were performed using data derived from tissue samples representing three cortical areas in three independent cohorts (ROSMAP, frontal; Knight ADRC, parietal; Mayo, temporal). C. Trans-eQTL analyses were performed in human microglia clusters from single nuclei RNA-seq. D. Functional genomics were applied to define the impact of rs1582763 and rs6591561 on microglia function using iTF-microglia. E. Summary of major findings.

**Figure 2.**
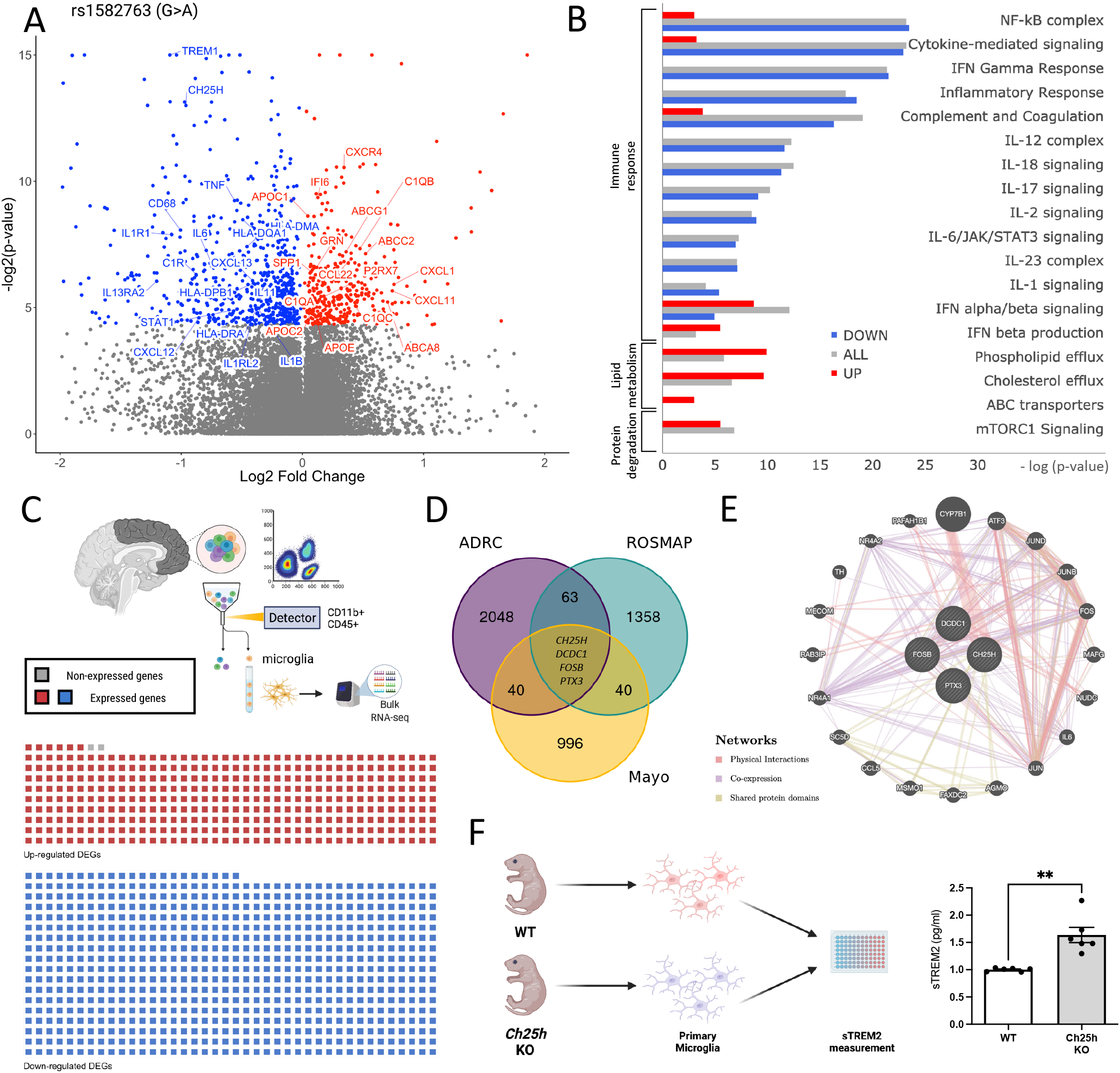
Brain transcriptomics reveals that the AD protective variant rs1582763 reduces the inflammatory response and increases lipid metabolism pathways. A. Meta-analysis of the impact of rs1582763 on gene expression in 3 independent brain cohorts. Volcano plot of genes that are differentially expressed in the presence of rs1582763. B. Pathway analysis reveals genes enriched in pathways related to immune signaling and lipid metabolism. Red, up-regulated genes. Blue, down-regulated genes. Gray, all differentially expressed genes. C. Sorted brain microglia (ROSMAP; n=10 brains) were queried to determine whether identified DEGs were enriched in microglia. The waffle plot illustrates the number of genes present in FACS-isolated microglia. Red, up-regulated genes. Blue, down-regulated genes. Gray, absent in the microglia dataset. D. Venn diagram shows the number of differentially expressed genes within three independent cohort (FDR < 0.05). Covariates include age, sex and disease status. E. Gene network diagram reveals the relatedness of *CH25H, DCDC1, FOSB*, and *PTX3*. Network: Red, physical interaction. Purple, co-expression. Olive, shared protein domain. F. Schematic of primary microglia culture from WT and *Ch25h* KO mice. Primary mouse microglia from *Ch25h* KO mice display significantly higher sTrem2 levels in the media detected by ELISA (p = 1.0 × 10^−3^).

Many genes differentially expressed based on rs1582763 genotype were known to be enriched in microglia, including *TREM1, CH25H, CXCR4, HLA* genes, *CD68*, among others. To determine whether these genes were statistically enriched in microglia expression, we analyzed transcriptomic data from Fluorescence-Activated Cell Sorting (FACS)-isolated microglia derived from human brains (n = 10; ROSMAP)^17,18^. Of the 1069 differentially expressed genes, 1067 were detected in the human brain-isolated microglia (Supplemental Table 1; Figure 2C). Together, these results suggest that rs1582763 primarily drives transcriptional changes in microglia.

Parallel trans-eQTL analyses were run within each of the brain cohorts. These more stringent analyses revealed an association between rs1582763 and reduced levels of *CH25H* (*β*=-0.37, p=1.22×10^−4^), *DCDC1* (*β*=-0.26, p=3.16×10^−5^), *PTX3* (*β*=-0.41, p=5.82×10^−5^), and FOSB (*β*=-0.49, p=4.38×10^−6^) (Figure 2D; Supplemental Table 5). These genes are involved in cholesterol metabolism and immune response and are highly related to one another by physical interaction, co-expression, and shared protein domains (Figure 2E). *DCDC1* is expressed in microglia^19^ and helps regulate microtubule polymerization and dynein-dependent transportation^20^. *PTX3* is predominately induced by pro-inflammatory cytokines and regulates complement activities in neuroinflammatory conditions such as multiple sclerosis (MS) and experimental autoimmune encephalomyelitis (EAE)^21-24^. FOSB was found to be expressed in activated microglia and plays a role in complement regulation^25^. *CH25H* encodes the microglia-specific enzyme, cholesterol 25-hydroxylase, and is involved in many of the key pathways altered by rs1582763. *CH25H* converts cholesterol into 25-hydroxysterol (25HC) which is considered an inflammatory mediator in microglial recruitment that promotes IL1B and NLRP3 inflammasome-mediated pro-inflammatory response^26^. Given that rs1582763 also results in elevated sTREM2 in human CSF ^10^, we sought to investigate whether *CH25H* modulates sTREM2 levels. Microglia isolated from *Ch25h* KO mice (postnatal day 2) produced significantly elevated sTrem2 compared to litter-mate controls (p= 1.0×10^−3^; Figure 2F). Thus, rs1582763, in the *MS4A* locus, regulates microglia-enriched genes, whose expression may impact sTREM2 levels.

### The protective variant impacts a specific population of microglia in human brains that regulates cytokines, chemokines, and the Type-I interferon pathway

Microglia are highly dynamic and context-dependent^27-30^. To further investigate the impact of rs1582763 on microglia states, we analyzed a cohort of AD and control brains with genomic and snRNA-seq data (n=66; 39% GG, 42% AG, 19% AA, Figure 1C; Figure 3A; Supplemental Table 1). Microglia were identified and annotated by cell-type specific markers (n=17,089)^31^ and grouped into nine distinct clusters (Mic.0-Mic.8) by performing unbiased clustering as previously described^32,33^. We then compared the microglial states associated with the minor and major alleles of rs1582763. The minor allele of rs1582763 was associated with a significant increase in the proportion of nuclei within the Mic.3 cluster (p=1.65×10^−3^; Figure 3B; Supplemental Figure 1; Supplemental Table 6). Within Mic.3, rs1582763 was associated with *MS4A4A* expression, whereby minor allele carriers exhibit significantly increased *MS4A4A* levels (p=1.11×10^−2^; Kruskal-Wallis test; Figure 3C). *MS4A4A* and *TREM2* expression was concentrated in Mic.3, while *MS4A6A* and *MS4A4E* expression patterns differed (Supplemental Figure 2). We observed the highest number of differentially expressed genes in Mic.3 (107 genes, FDR<0.05; Figure 3D). Thus, the primary molecular effect of rs1582763 occurred within a specific microglia population: Mic.3.

**Figure 3.**
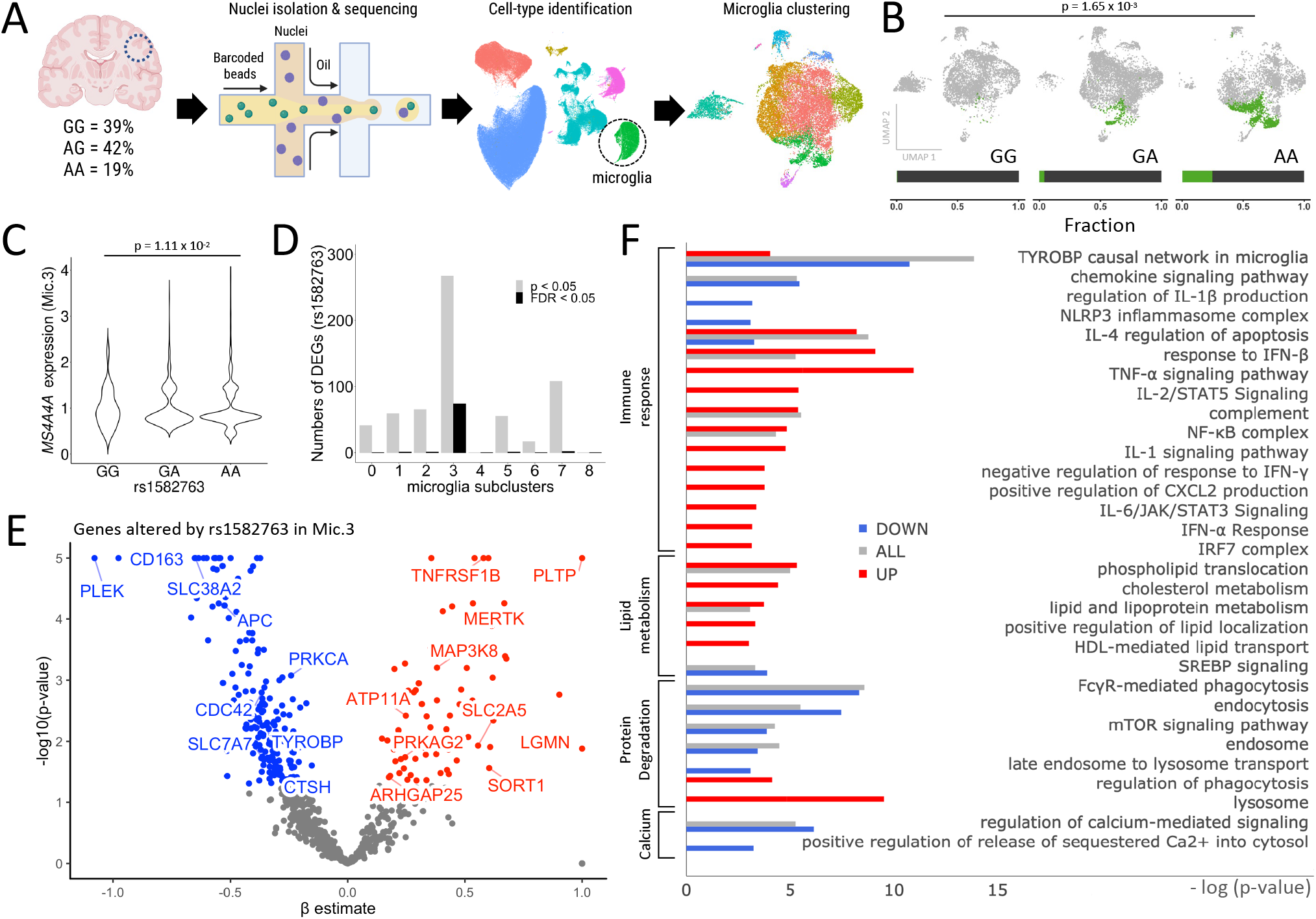
Single nucleus RNA-seq in human brains reveals that the AD protective variant rs1582763 alters a specific microglia population and drives the expression of genes associated with chemokine and lipid metabolism pathways. A. Schematic of single nucleus brain RNA-seq. B. UMAP reveals a significant increase in the proportion of a specific population of microglia (Mic.3) in the presence of the rs1582763 minor allele (A) (The fraction below indicates the proportion of Mic.3 in the total microglia of given genotype carriers). C. *MS4A4A* expression is significantly elevated in Mic.3 in the presence of the minor allele of rs1582763 (Kruskal-Wallis test, p = 1.11 × 10^−2^). D. Bar graph of the number of genes differentially expressed as a function of the rs1582763 genotype across microglia subclusters reveals that differentially expressed genes are enriched in Mic.3. Grey, p<0.05. Black, FDR<0.05. E. Volcano plot of genes differentially expressed based on rs1582763 genotype in Mic.3. F. Pathway analysis reveals that rs1582763 alters genes within Mic.3 that influence immune response, lipid metabolism, protein degradation and calcium regulation.

Given the impact of rs1582763 on gene expression specifically in Mic.3, we sought to define the genes that were changed as a function of rs1582763 within this subpopulation of microglia. We identified 259 genes that were significantly differentially expressed within Mic.3 according to rs1582763 genotype (Figure 3E). Many of these genes influence pathways related to immune function (TYROBP [p=2.1×10^−5^], Chemokine [p = 8.39×10^−3^], TNF-⍰ [p = 3.65×10^−3^], NLRP3 inflammasome [4.54×10^−2^]) and are associated with an increased Type-I interferon response (IFN-β [p = 1.11×10^−4^]). Other genes are also enriched in pathways related to enhanced lipid metabolism (p = 2.39×10^−2^) enhanced protein degradation (mTOR signaling [p = 2.09×10^−2^], lysosome [p = 9.11×10^−3^]), and decreased calcium regulation ([p = 4.31×10^−3^]) (Figure 3F; Supplemental Table 8).

Together, we showed that rs1582763 mediates reduced inflammatory gene expression and enhanced cholesterol metabolism gene expression in a specific population of microglia in the human brain. These pathways were also highly related to TREM2 function, and in primary microglia, targeting one of these genes, *CH25H*, was sufficient to shift sTREM2 levels in a manner consistent with the protective variant.

### AD risk variant MS4A4A rs6591561 promotes inflammasome response and increases intracellular cholesterol storage

To understand the impact of the variant in *MS4A4A* that is associated with increased AD risk (rs6591561), we evaluated gene expression patterns in AD brains among rs6591561 variant carriers (AA, GA, GG; Figure 1A-B). Meta-analysis of three brain cohorts (n=579 brains in Knight ADRC, Mayo, and ROSMAP; Figure 4A; Supplemental Table 1) revealed 1597 differentially expressed genes as a function of the rs6591561 genotype (FDR<0.05; Supplemental Table 9) including *MS4A4A* (p = 3.8×10^−2^), which showed reduced expression. These genes were also enriched in sorted microglia from human brains (1593/1597; Figure 4B). Genes altered by rs6591561 are involved in pathways associated with elevated pro-inflammatory responses (NF-kB [p=1.04×10^−6^], cytokine-mediated signaling [p=2.75×10^−7^], IFN-γ [p=1.78×10^−6^]); reduced Type-I interferon response (IL-4 [p=4.61×10^−7^], IL-10 [p=1.92×10^−3^], IFN-α/β[p=1.59×10^−6^]); and reduced lipid metabolism (cholesterol efflux [p = 5×10^−3^]) (Figure 4C; Supplemental Table 10). Interestingly, many of these pathways were shared with the protective variant but with opposing effects. Several genes in the AD risk loci, including *MS4A4A*, were significantly reduced among minor allele carriers: *HLA-DRA, HLA-DQA1, MS4A4A, HLA-DPB1, CFD, C2, PVRIG, PCOLCE, TREM1, HLA-DMA, MIDN, AQP9, MBLAC1, HLA-DOA, KCNJ13, AZU1*, and *CYP27C1* (Supplemental Table 11).

**Figure 4.**
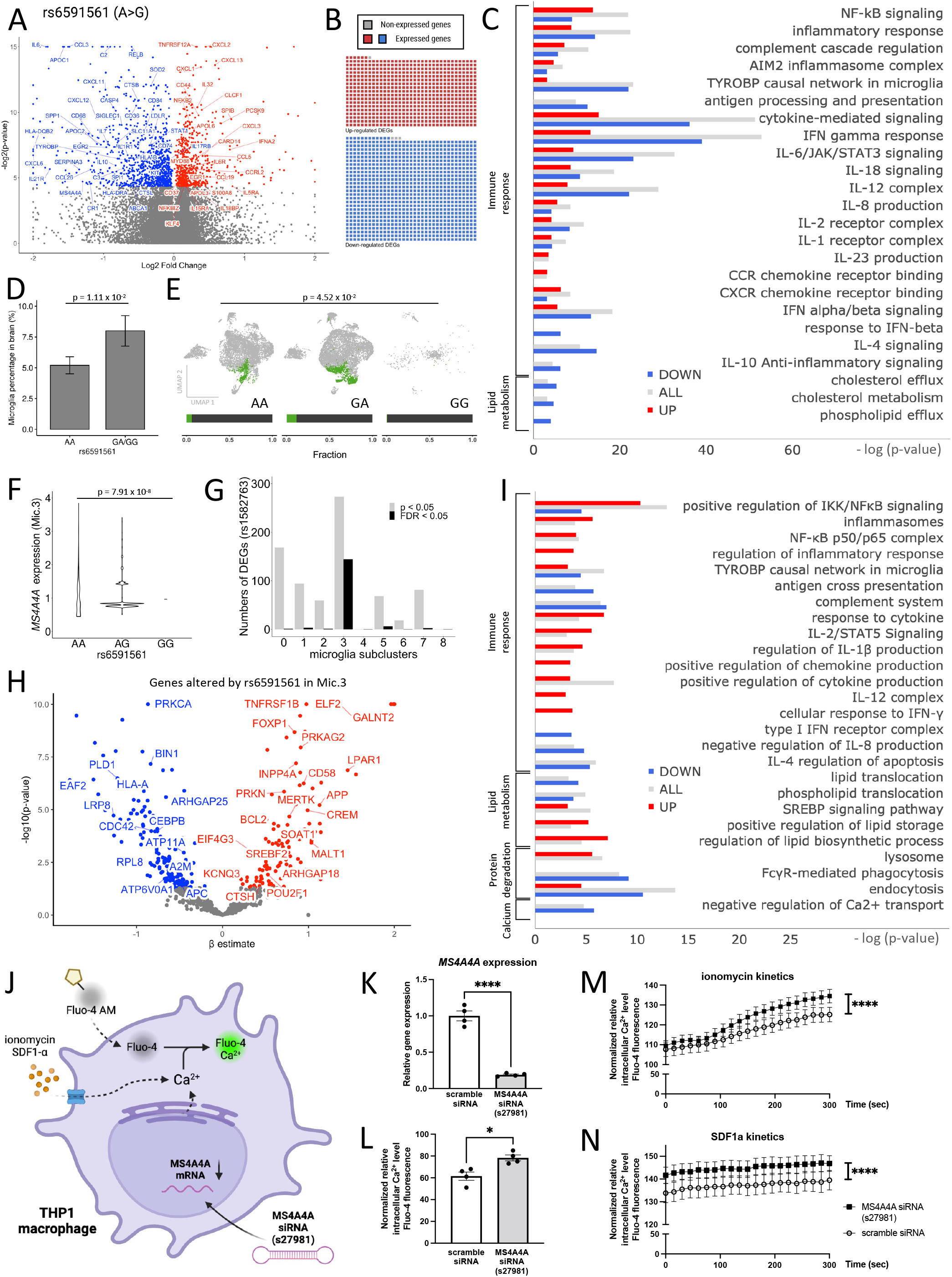
The AD risk variant rs6591561 is associated with upregulation of inflammasome and inflammatory cytokines and reduced lipid metabolism. A. Meta-analysis of the impact of rs6591561 on gene expression in three independent brain cohorts. Volcano plot of genes that are differentially expressed in the presence of rs6591561. B. Sorted brain microglia (ROSMAP; n=10 brains) were queried to determine whether genes identified in (A) were enriched in microglia. The waffle plot illustrates the number of genes present in sorted microglia. Red, up-regulated genes. Blue, down-regulated genes. Gray, absent in the microglia dataset. C. Pathway analysis of genes from (A) reveals genes enriched in pathways related to immune signaling and lipid metabolism. Red, up-regulated genes. Blue, down-regulated genes. Gray, all differentially expressed genes. D. Single nucleus RNA-seq reveals that rs6591561 is associated with an increased microglia proportion (LR, p = 1.11 × 10^−2^). E. UMAP reveals a significant increase in the proportion of a specific microglia population of microglia (Mic.3) in the presence of the minor allele of rs6591561 (LR, p = 4.52 × 10^−2^). F. *MS4A4A* expression is reduced in minor allele carriers of rs6591561 in Mic.3 (ANOVA, p = 7.91 × 10^−8^). G. Bar graph of the number of genes differentially expressed as a function of the rs6591561 genotype across microglia subpopulations reveals that differentially expressed genes are enriched in Mic.3. Gray, p<0.05. Black, FDR<0.05. H. Volcano plot reveals genes differentially expressed based on the rs6591561 genotype in Mic.3. I. Pathway analysis reveals that rs6591561 alters genes involved in elevated inflammasome, inflammatory cytokines, enhanced lipid storage, and decreased lipid metabolism in Mic.3. J. Schematic representation of Fluo-4 calcium assay on THP-1 macrophage treated with *MS4A4A* siRNA (s27981). K. qPCR reveals *MS4A4A* is significantly reduced in the presence of *MS4A4A* siRNA. L. Baseline intracellular Ca^2+^ levels are elevated after treatment with *MS4A4A* siRNA s27981. M. Intracellular Ca2+ kinetics after treated with SDF1α. N. Intracellular Ca2+ kinetics after treated with ionomycin ****, p<0.0001.

Next, we examined the impact of rs6591561 on specific microglia states using snRNA-seq data from human brains. We detected a significant increase in total microglia nuclei among carriers of the minor allele in rs6591561 (p=1.11×10^−2^; Figure 4D). Among microglia subclusters, nuclei in Mic.3 were significantly increased in heterozygous carriers of rs6591561 in comparison to those homozygous for the major allele (p = 4.52×10^−2^; Figure 4E). Consistent with the eQTL between rs6591561 and *MS4A4A* observed in brain homogenates (Figure 4A), we detected a significant eQTL between rs6591561 and *MS4A4A* in Mic.3 (Figure 4F; Supplemental Figure 1). Mic.3 also exhibited the highest number of differentially expressed genes as a function of the rs6591561 genotype (90 genes, FDR<0.05; Figure 4G; Supplemental Table 12). Thus, rs6591561 drives molecular changes in a specific microglia state: Mic.3.

Genes differentially expressed in Mic.3 due to rs6591561 were associated with immune response (inflammasomes [p=3.53×10^−3^], NF-kB [p=3.33×10^−5^], cytokine [p=1.17×10^−3^], IL-2/STAT5 [p=4.01×10^−3^]), cholesterol metabolism (lipid storage [p=5.37×10^−3^]), protein degradation (Lysosome [p=4.65×10^−3^]), and calcium regulation (negative regulation of Ca2+ transport [p=3.10×10^−3^], Figure 4H-I; Supplemental Table 13). Calcium homeostasis is critical to the function of many of the pathways identified. To test whether *MS4A4A* expression impacts intracellular calcium levels, we silenced *MS4A4A* with RNAi in THP-1 myeloid lineage cells. Calcium levels were monitored by Fluo-4 AM calcium indicator (Figure 4J). Consistent with the snRNA-seq findings, silencing *MS4A4A* expression produced significantly higher intracellular Ca^2+^ levels at baseline compared with scrambled controls (p=0.01; Figure 4L). Additionally, increased release of intracellular Ca^2+^ stores with ionomycin led to a significant increase in Ca^2+^ levels when *MS4A4A* was silenced (Figure 4M). In the presence of a calcium effector, CXCL12 (SDF-1α, a ligand for CXCR4), silencing *MS4A4A* led to elevated intracellular Ca^2+^ kinetics compared to scrambled-treated cells (Figure 4N). These data suggest a role for *MS4A4A* in regulation of calcium homeostasis in microglia. Together, we find that rs6591561 induced molecular and functional effects in microglia in human brains.

### Microglia cluster 3 is a chemokine/cytokine-specific cluster

Microglia exhibit tremendous transcriptional diversity in human brains, which may reflect dynamic changes due to aging, environment, and disease states ^34^. Microglia subclusters were defined using unsupervised clustering^32^. Transcriptionally, Mic.0 was enriched in genes associated with homeostatic states (*SELPLG, MED12L, BIN1, CX3CR1, P2RY13, TMEM119*), while Mic.1 was enriched in genes found in activated and disease-associated microglia states (*CD83, TNFAIP3, C5AR1, CD68, GPNMB, ABCA1*). However, the identity of Mic.3, which was impacted by both protective and risk variants, was unclear.

To begin to understand the identity of this microglia state, we compared gene expression in Mic.3 to other the microglia subclusters in the brain (p < 0.05; Figure 1D; Figure 5A). We identified more than 150 genes within Mic.3 that were significantly upregulated, including several AD risk genes: *BIN1* (p=8.58×10^−12^), *HLA-DRB1* (p=1.27×10^−2^), *SLC12A9* (p=2.67×10^−4^) (Figure 5A-5B; Supplemental Table 14). Mic.3-specific genes are enriched in pathways associated with elevated immune response, such as cytokine and chemokine pathways (IL-2 [p=1.18×10^−12^], NF-kB [p=1.4×10^−2^], CXCR4 [p=1.16×10^−2^], IFN-γ [p=8.85×10^−6^]), and other pathways like TLR [p=1.33×10^−6^], TNF-a [p=8.64×10^−6^], and TGF-*β* [p=1.33×10^−2^] (Figure 5B; Supplemental Table 15). Additionally, while *CH25H* was significantly elevated, other lipid metabolism genes were reduced (p=3.28×10^−2^) (Figure 5B; Supplemental Table 15).

**Figure 5.**
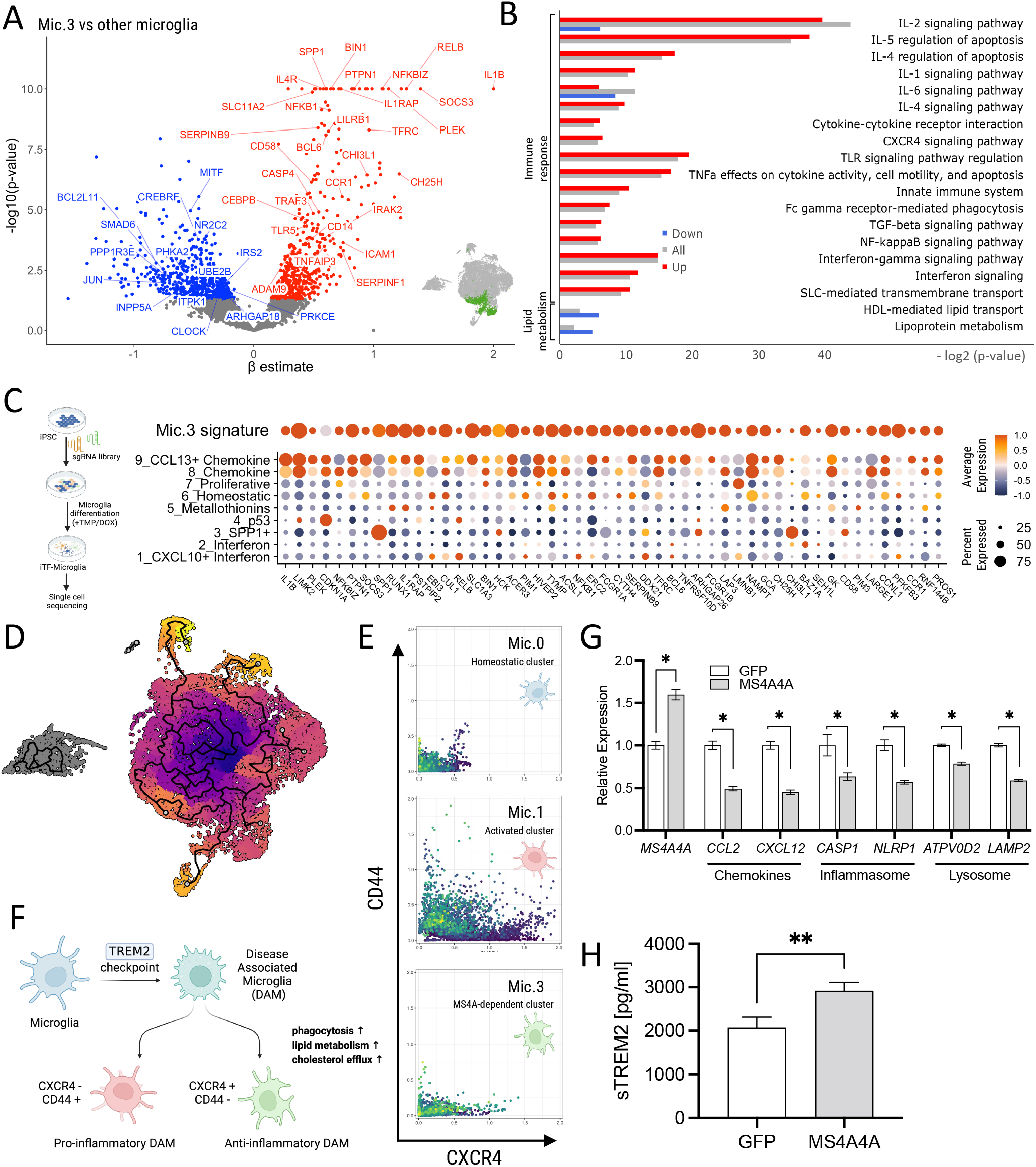
Variants in the MS4A locus shift microglia identity. A. A volcano plot illustrates differentially expressed genes between Mic.3 and all microglia subclusters. The inset graphic shows the reference location of Mic.3. B. Pathway analysis reveals enrichment of genes associated with chemokines, immune/inflammatory response and lipid metabolism. Red, upregulated DEGs. Blue, downregulated DEGs. Gray, all differentially expressed genes. C. (Left) A graphic illustration shows CROP-seq from iTF-microglia. (Right) A dot plot shows the expression level of the top 50 Mic.3 differentially expressed genes in Knight ADRC brain microglia and iTF-microglia. Average expression indicated by color scale. Percent expressed illustrated by the size of the circle. D. An UMAP diagram shows homeostatic (Mic.0), activated (Mic.1) and MS4A4A-dependent (Mic.3) microglia clusters trajectory by Monocle 3. E. Scatterplots show imputed relative expression of CXCR4 (anti-inflammatory)/CD44 (pro-inflammatory) in homeostatic (Mic.0), activated (Mic.1), and MS4A4A-dependent (Mic.3) microglia cluster generated by MAGIC. F. A hypothesis proposes that disease-associated microglia (DAM) differentiate into the pro-inflammatory DAM and the anti-inflammatory DAM. G-H. Peripheral blood mononuclear cell (PBMC) derived macrophages were transduced with lentivirus particles carrying GFP or MS4A4A WT. G. RNA expression levels of *MS4A4A*, chemokine, inflammasome, and lysosomal genes were plotted. H. sTREM2 levels in media from GFP or *MS4A4A*-encoding lentiviral transduced macrophages. *, p < 0.05; **, p < 0.001.

Due to the pleiotropic nature of cytokines and chemokines, the expression patterns of these genes and pathways alone were not sufficient to clarify the identity of Mic.3. To refine the identity of Mic.3, we leveraged a recently described dataset from human microglia-like cells (induced transcription factor (iTF)-microglia) as an *in vitro* reference. iTF-microglia were transformed into a spectrum of transcriptionally diverse microglia via CRISPRi-based editing^35^. The top 50 differentially expressed genes that define Mic.3 were set as a gene module and visualized by calculating the average expression score of the iTF-microglia dataset on a single-cell level. Genes defining Mic.3 were enriched in iTF-microglia cluster 8 and cluster 9, which were associated with chemokine and C-C motif chemokine ligand 13 (CCL13+) chemokine functions, respectively (Figure 5C).

### Mic.3 contains anti-inflammatory microglia that are regulated by MS4A4A expression

Microglia exhibit multiple activation states in response to distinct contexts. Microglia transition to a TREM2-dependent disease-associated microglia (DAM) activation state that can be further sub-classified into pro-inflammatory DAM (*CD44+*) and anti-inflammatory DAM (*CXCR4+*) states^36^ (Figure 5D). Mic.3 shares many activation markers with Mic.1, suggesting Mic.1 and Mic.3 may both represent sub-types of activated microglia in the human brain^32^.

To investigate this possibility, we evaluated whether homeostatic microglia (Mic.0) were equally likely to transition to the populations of activated microglia (Mic.1 and Mic.3). We employed pseudo-time analytical frameworks, including Monocle3^37^ and velocyto^38^, to predict the transcriptomic trajectories across Mic.0, Mic.1, and Mic.3 (Figure 5E). We found that Mic.0 points outward, then diverges in opposite directions toward both Mic.1 and Mic.3 (Supplemental Figure 3). Thus, we hypothesized that Mic.1 and Mic.3 represent pro-inflammatory (*CD44+*) and anti-inflammatory (*CXCR4+*) DAM states. High-dimensional geometry diffusion analysis was performed on *CD44/CXCR4* expression levels in each cluster using Markov Affinity-based Graph Imputation of Cells (MAGIC)^39^. Mic.0 expressed *CD44* and *CXCR4* at low levels, consistent with a homeostatic state. Mic.1 exhibited high *CD44* and *CXCR4*, suggesting equal proportions of pro- and anti-inflammatory DAMs. Interestingly, Mic.3 nuclei were high in *CXCR4*, while *CD44* remained low, consistent with an anti-inflammatory-enriched pool (Figure 5F; Supplemental Figure 4). *CXCR4+* anti-inflammatory DAM has been reported to exhibit enhanced phagocytosis, lipid metabolism, and cholesterol efflux^36^.

Many of these features are also found in rs1582763 carriers, leading us to hypothesize that *MS4A4A* expression regulates microglial chemokine/cytokine function and promotes an anti-inflammatory DAM cell state. To test this hypothesis, human peripheral blood mononuclear cells (PBMCs) were transduced with Lentiviral particles containing *MS4A4A* or GFP, mimicking the effect of rs1582763 on *MS4A4A*. Overexpression of *MS4A4A* led to a reduction in chemokine, inflammasome, and lysosome genes (Figure 5G) along with an increase in sTREM2 levels (Figure 5H). This result was consistent with the observation that rs1582763 carriers have higher sTREM2 levels in CSF^10^ and reduced expression of genes associated with chemokine, inflammasome, and lysosome pathways in human brain tissue samples (Figure 3F).

### MS4A4A protective variant rs1582763 modulates microglial behavior by inhibiting chemokine response and promoting interferon response

It was surprising to identify a single microglia state that was altered by variants with opposing effects on AD risk in the *MS4A* gene locus. To test how protective and risk variants impact the microglia population, Mic.3, we leveraged the CROP-seq data from iTF-microglia, which produced a spectrum of microglial phenotypes from “interferon” to “chemokine” by silencing genes with a druggable sgRNA library ^35^. Interestingly, *MS4A4A* expression was enriched in the interferon cluster (Figure 6A), and inversely correlated with the expression of the genes enriched in Mic.3, which mapped to the chemokine clusters in the iTF-microglia (Figure 6A).

**Figure 6.**
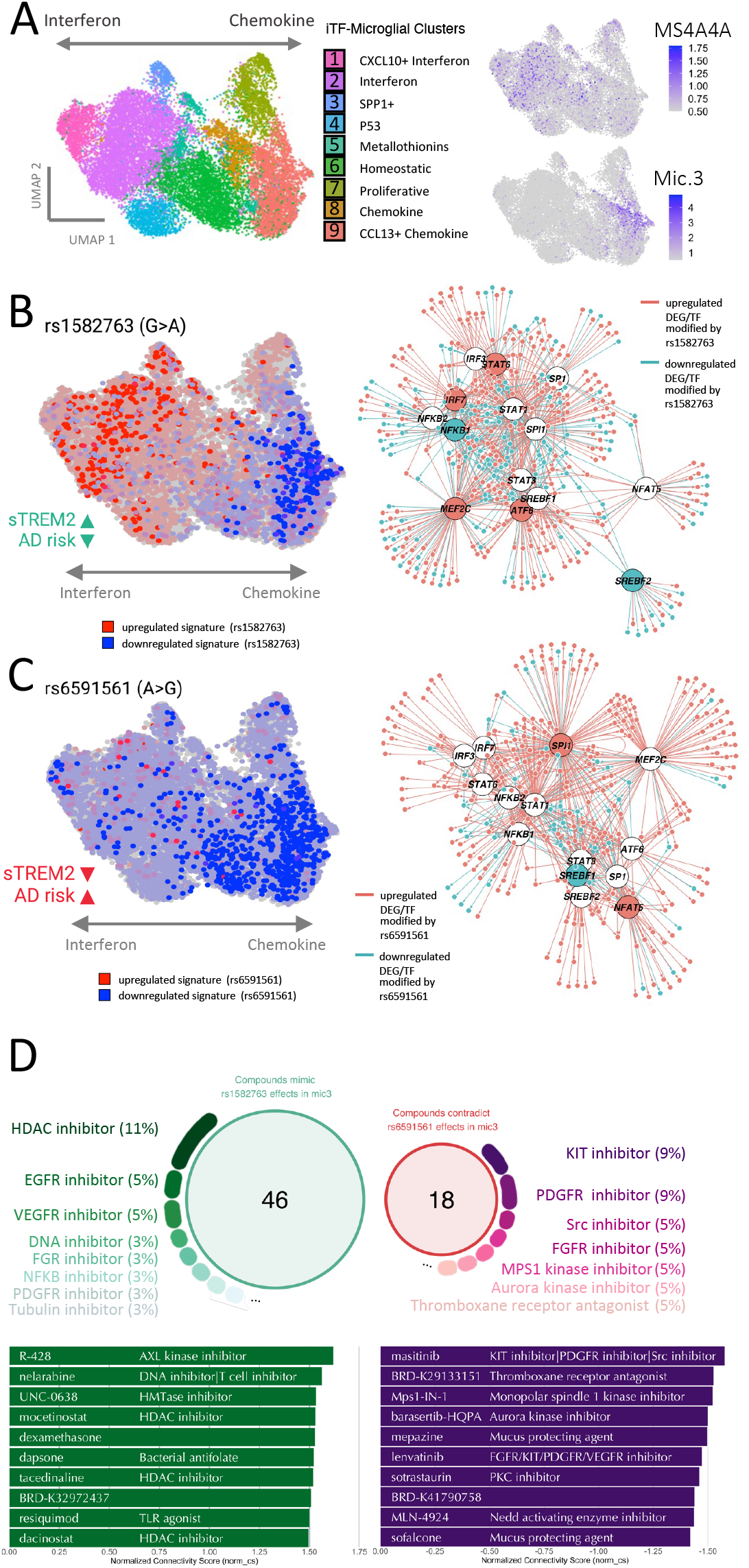
The AD protective and risk variants in the MS4A locus alters the microglial state transcriptionally and functionally. A. Left, schematic of CROP-seq from iTF-microglia. Right, *MS4A4A* expression is enriched in the iTF-microglia cluster associated with interferon function (top). Right, genes differentially expressed in Mic.3 compared to all microglia subclusters are enriched in iTF-microglia chemokine clusters (bottom). B. Molecular signature of the protective variant is enriched in interferon iTF-microglia (left) and alters transcription factor networks (right). C. Risk variant signature is dampened across interferon and chemokine iTF-microglia (left) and alters transcription factor networks (right). D. Compounds that mimic the molecular signature of the protective and reverse risk variant effects in Mic.3 are shown along with the target percentage and top 10 compounds.

To determine how the protective variant impacts microglia identity, we mapped the genes that were associated with rs1582763 genotype in Mic.3 on the iTF-microglia (Figure 6B). We found that the upregulated genes, including *MS4A4A*, were enriched in iTF-microglia clusters associated with interferon signaling (Figure 6B), while the downregulated genes were localized to the chemokine clusters. Thus, the AD-protective variant may regulate Mic.3 by promoting interferon function and inhibiting chemokine function.

Interestingly, genes that were dysregulated as a function of the rs6591561 genotype in Mic.3 were mapped to the chemokine cluster in the iTF-microglia (Figure 6C). The mapping of rs6591561-associated genes suggested an overall repression of the chemokine clusters that define Mic.3 in conjunction with a failure to promote the interferon function (Figure 6C).

Given the evidence of the regulatory potential of rs1582763 and rs6591561, we sought to determine whether genes enriched in Mic.3 are targets of specific transcription factors using ChEA3, a web-based transcription factor enrichment analysis application^40^. Mic.3 genes were regulated by transcription factors enriched for microglial (*SPI1, MEF2C*), chemokine (*NFKB1, NFKB2*), interferon (*STAT1, STAT3, STAT6, IRF7*), and lipid metabolism (*SREBF1, SREBF2*). The AD protective variant, rs1582763, altered genes associated transcription factors that drive the upregulation of interferon (*IRF7* and *STAT6*), downregulation of chemokine (NFKB1), and downregulation of cholesterol synthesis (*SREBF2*) (Figure 6B, right). Conversely, the AD risk variant, rs6591561, was associated the upregulation of microglial transcription factors (*SPI1*) and downregulation of lipid metabolism transcription factors (*SREBF1*) (Figure 6C, right). SPI1 has been reported to be an AD risk factor^1,41,42^.

### Leveraging FDA-approved drugs to promote protective effects of MS4A4A

To better understand the impact of the *MS4A4A* variants and leverage their therapeutic potential, we sought to identify the compounds that mimic the gene expression changes observed in the protective and risk variants. To achieve this, we used Connectivity Map (CMap)^43,44^ to search for drug candidates with effects that mimic the molecular signatures associated with each variant. The CMap database contains over one million gene expression signatures from multiple cell lines treated with more than 80K perturbagens. We focused on findings from hemopoietic cell lines (BJAB, CD34, HAP1, HBL1, HL60, JURKAT, K562, KMS34, MINO, NALM6, NOMO1, OCILY10, OCILY19, OCILY3, THP1, TMD8, U937) treated with compound perturbagens. We identified 46 compounds (norm_cs > 1.35) that uniquely mimic the protective variant gene signature (Figure 6D). These compounds were enriched in HDAC inhibitors (11%). We also identified 18 compounds that reverse the gene signature created by the risk variant. These compounds were enriched in KIT inhibitors (9%) and PDGFR inhibitors (9%) (Figure 6D). In parallel, we identified 14 drug candidates that reverse the molecular signatures produced by the protective variant and 75 drug candidates that mimic the gene signatures produced by the risk variant. (Supplemental Figure 5). Thus, we identified compounds that promote the protective effects of rs1582763 and counteract the detrimental effects of rs6591561 for consideration in therapeutic design.

## Discussion

Variants in the *MS4A4A* locus are associated with age-at-onset of AD and AD risk; however, the role of *MS4A4A* in AD pathogenesis remains poorly understood^1-5^. Our study shows that the AD protective variant drives elevated *MS4A4A* expression and shifts the chemokine microglia subpopulation to a more interferon state. In contrast, the AD risk variant suppresses *MS4A4A* expression and diminishes the chemokine microglia subpopulation with an inability to promote the interferon state (Figure 1E). We also show that we can leverage these findings for their therapeutic potential by predicting drugs that recapitulate the protective variant molecular signatures or reverse the risk variant molecular signatures.

Chemokines, cytokines, and their receptors modulate cellular events such as chemotaxis, phagocytosis, cytokine secretion, cell activation, proliferation, survival, and death. As a result, the dysregulation of chemokines or cytokines play a critical role in neuroinflammation and neurodegeneration. Under steady-state conditions, homeostatic chemokines and cytokines are involved in immune surveillance and modulation of immune cell differentiation during hematopoiesis. Conversely, proinflammatory chemokines and cytokines activate and recruit microglia and release more inflammatory mediators in the disease state. Occasionally, persistently activated microglia lead to chronic neuroinflammation, which may exacerbate AD. *MS4A4A* is reportedly a surface marker for the anti-inflammatory M2 macrophage^45^. Here, we show that rs1582763 increases *MS4A4A* expression and decreases the inflammatory cytokine and chemokine responses in both bulk and snRNA-seq. Specifically, pro-inflammatory chemokines like CCL3 and CCL4 decrease in bulk and snRNA-seq, while anti-inflammatory IL4 receptors are upregulated. Furthermore, carrying the minor allele of rs1582763 directly correlates with the emergence of a specific microglia cluster (Mic.3), which is a *MS4A4A*-dominant cluster that persists in an anti-inflammatory DAM state (CXCR4 high/CD44 low)^36^. Conversely, rs6591561 results in decreased *MS4A4A* expression and elevated inflammasome and inflammatory responses. These results implicate a direct link between the increased *MS4A4A* expression and the decreased inflammatory response in microglia.

Cholesterol dyshomeostasis is a major risk factor for AD^46^. Beyond APOE4 ^47-49^, growing evidence supports the role of intracellular cholesterol metabolism in microgliosis. Specifically, the accumulation of intracellular cholesterol esters (CE) impedes physiological immune surveillance and scavenger function, promotes chronic inflammatory activation, and ultimately leads to inflammasome activation in microglia ^50^. TREM2 also plays a critical role in cholesterol metabolism. TREM2 is required for DAM activation^28^. TREM2 shifts the energy state by promoting glycolysis and mTOR signaling^51,52^. TREM2 also activates the LXR transcriptional factor and initiates ABC transporters to promote cholesterol efflux, thus preventing lysosomal dysfunction^53^. Our data suggest that *MS4A4A* variants rs1582763 and rs6591561 regulate lipid metabolism, such that rs1582763 promotes ABC transporter expression and cholesterol efflux, while rs6591561 downregulates the ABC transporters and promotes cholesterol storage. The AD protective variant, rs1582763, upregulates *LRP* (ApoE, Tau, and Aβ receptor), *PLTP* (converting VLDL to HDL), and *SORT1* (VLDL transportation). Alternatively, the AD risk variant, rs6591561, upregulates *PCSK9* (LDLR degradation) and *SOAT* (promoting CE storage). CSF PCSK9 is elevated in AD patients, and PCSK9 expression is associated with AD pathogenesis^54,55^. A PCSK9 inhibitor reduces neuronal inflammation and Aβ aggregation in rodents^56^. These findings suggest *MS4A4A* expression stymies AD progression by sustaining cholesterol homeostasis in the brain. MS4A4A and TREM2 co-localize in lipid rafts ^57^. This result, to our knowledge, is the first evidence that connects MS4A4A expression with cholesterol metabolism in microglia, suggesting that MS4A4A facilitates cholesterol efflux in microglia.

*CH25H* was found to be significantly downregulated among protective variant carriers (rs1582763), suggesting a linkage between elevated *MS4A4A* expression and reduced *CH25H* expression. Furthermore, cultured microglia with reduced *Ch25h* expression shed more sTrem2 into culture media. The product of *CH25H*, 25-hydroxycholesterol (25-HC), is one of the major microglia-secreted inflammatory mediators in the brain^58-61^. Previous studies have shown that *CH25H* plays a pivotal role in immune response and cholesterol metabolism. For instance, *CH25H* is one of the downstream members of TREM2-mediated lipid metabolism^53^ and takes part in cholesterol biosynthesis via the SREBP signaling pathway^62^. The product of Ch25h enzyme activity, 25-hydrocholesterol (25-HC), is a well-studied suppressor of cholesterol biosynthesis via the SREBP2 pathway ^63,64^. Both anti-inflammatory^65,66^ as well as proinflammatory^26,59^ activities have been reported for 25-HC. In the CNS, 25-HC produced by activated microglia appears to have proinflammatory effects^67,68^. Our data show that decreased *Ch25h* levels leads to increased sTrem2 levels and an interferon response phenotype pf microglia that may be beneficial. These results point to *CH25H* as a mediator by which *MS4A4A* to regulates immune response and cholesterol metabolism in AD pathophysiology.

The transcriptional effects of *MS4A4A* variants, rs1582763 and rs6591561, were divergent; however, their effects were localized to the same microglia subpopulation. Microglia subcluster 3 (Mic.3) was altered specifically by both variants. This cluster may be highly relevant to AD pathophysiology as several top upregulated genes, such as *BIN1, HLA-DRB1*, and *SLC12A9*, were positively correlated with AD susceptibility^4^. Mic.3 likely plays a role in chemokine/chemokine CCL13+ function and is functionally distinct from the homeostatic and activated microglia that also exist in the AD brains^33^. Mic.3 likely acts as an alternative activation state that is enriched in Type-I interferon function.

In summary, we reported bidirectional biological effects among two variants located in the *MS4A4A* locus. The AD protective variant, rs1582763, featured higher CSF sTREM2 and lower AD risk, was found to have increased *MS4A4A* expression, lower chemokine and cytokine immune responses, and higher cholesterol efflux (Figure 1E). The AD risk variant, rs6591561, featured lower CSF sTREM2 and higher AD risk, was found to have decreased *MS4A4A* expression, and greater expression of genes associated higher inflammatory cytokines, chemokines, and the inflammasome responses, as well as higher cholesterol storage (Figure 1E). Combining the genotypic and brain transcriptomic data from large AD and control cohorts, our study provides a possible mechanistic explanation for how variants in *MS4A* loci alter the AD risk and the underlying causes for how *MS4A4A* modifies immune response and cholesterol metabolism in AD pathophysiology.

## Online Methods

### Bulk and single nucleus RNA-seq datasets

Bulk RNA-seq datasets were obtained from ROSMAP^69,70^, Mayo^71^, and DIAN/Knight ADRC^72^. The ROSMAP (Synapse ID = Syn3219045) and Mayo (Synapse ID = Syn5550404) datasets were downloaded from AMP-AD Knowledge Portal (https://adknowledgeportal.org). Briefly, 579 samples from neuropathologically confirmed AD (N = 400) and neuropathology-free controls (N = 179) were selected for differential expression and composition analyses. The cohort sample size, brain region, average age, sex percentage, allele frequency for the risk and protective variants, and disease status are summarized in Supplemental Table 1.

Bulk RNA-seq datasets for FACS-isolated microglia from ROSMAP were downloaded from AMP-AD Knowledge Portal and processed as previously described^32^. Ten samples, AD (N = 4) and healthy control (N = 6), were included in the analysis. Microglia were collected and sequenced. The sample size, brain region, age, sex, postmortem interval (PMI) and disease status are summarized in Supplemental Table 1.

The snRNA-seq dataset was obtained from Knight ADRC and processed as previously described^32^. Briefly, snRNA-seq of microglia from 66 samples with neuropathologically confirmed AD or neuropathology-free controls were used in the differential expression and compositional analyses. The sample size, brain region, age, sex, PMI, allele frequency for the risk and protective variants, and the disease status description are summarized in Supplemental Table 1.

Genetic data obtained from each study was used to determine the genotype of the risk and protective variants. The meta-data acquired from each study, for example, age, sex, PMI, RIN, TIN, and diagnosis/symptom, were used as covariates to define differential gene expression and identify compositional changes.

## Data analyses

### Functional analysis of bulk RNA-seq

Primary Differential Expression Gene (DEG) discovery in ROSMAP, Mayo, and Knight ADRC were identified using *DESeq2*^73^. To find DEGs for AD risk and protective variants, the age, sex, and disease status of the cohorts were used as covariates, and an additive model in the Likelihood Ratio Test (LRT) method was used for the variants. The overlap of DEGs

(threshold was set at *p*-value <0.05) between ROSMAP, Mayo, and Knight ADRC were cross-compared and shown in a Venn Diagram. The *p*-value and Log2 Fold Change (Log2FC) listed in the table were obtained from *DESeq2*. Four DEGs with a common direction across three datasets were put into GeneMania^74^ to define associated genes. The following formula design was used in *DESeq2*:

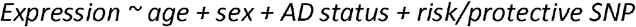

Meta-analysis was applied to combine the three independent brain cohorts using *MetaVolcanoR*. The Random Effects Model (REM) *rem_mv* was used to integrate *p*-values from *DESeq2*. Integrated *p*-values for candidate genes were ranked in ascending order, and the threshold was set at 5%. The top 5% ranked genes were identified as significant DEGs and plotted in a volcano plot.

DEGs identified by the meta-analysis were explored in FACS-isolated microglia from human brains. The transcripts per million (TPM) of sequenced genes from 10 brain samples were calculated and averaged. The average TPM for queried DEGs were summarized. Genes with an average TPM value greater than zero were considered expressed. The main figures show protein coding DEGs.

Pathway analysis was applied to DEGs using *EnrichR*^75^. Pseudogenes and non-coding transcripts were identified by ToppGene^76^ and removed before the query. Pathway enrichments were collected from “BioPlanet,” “KEGG human,” and “WikiPathway” gene sets.

The KEGG-based pathway integration and visualization tool *Pathview*^77^ was applied. The DEGs and REM-estimated summary fold-change (randomSummary) derived from the meta-analysis were diagrammed in the pathway ‘Cytokine-cytokine receptor interaction’ and ‘Cholesterol metabolism’ (Supplemental Figure 6).

### Functional analysis of single-nucleus RNA-seq

The processed, identified, and clustered single nuclei from microglia were used in this analysis. The detailed procedure can be found in a previous publication^32^. Briefly, microglia were clustered into nine expression states. For each expression state, the linear mixed model was applied to identify differentially upregulated genes, and those genes were used in a pathway analysis. Each expression state was associated with 12 previously described microglia functional states by hypergeometric test, to identify feature cell states like ‘homeostatic’ and ‘disease-associated’ microglia clusters.

Each microglia cluster was tested for DEGs associated with the risk or protective variants. The nuclei expressing 80% of all tested genes and the candidate genes expressed in 80% of nuclei were tested in each microglia cluster. The generalized linear mixed model (GLMM) *glmmTMB* ^78^ was used to test the DEGs for risk and protective variants, and age, sex, disease status. The alternative risk/protective variant was included among covariates. In each microglia cluster, the numbers of DEGs that passed the *p*-value and BH-FDR<0.05 threshold were recorded and plotted into a histogram. The following formula design was used:

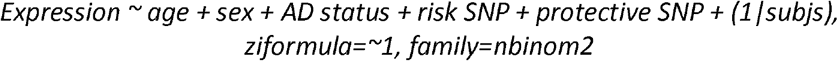

The DEGs for the risk/protective variant in Microglia cluster 3 (Mic.3) were then plotted in a volcano plot and examined more thoroughly for pathway analyses. Pathway analyses were applied to DEGs with *EnrichR*^75^. DEGs with a *p*-value < 0.05 were analyzed. Pseudogenes and non-coding genes were excluded before analysis. Pathway enrichments were collected from “BioPlanet_2019”, “WikiPathway_2021_Human”, “KEGG_2021_Human”, “GO_Biological_Process_2021”, “GO_Molecular_Function_2021”, “MSigDB_Hallmark_2020”, and “Jensen_COMPARTMENTS” databases.

The KEGG-based pathway integration and visualization tool *Pathview*^77^ was applied. The *p*-value and fold-change derived from GLMM *glmmTMB*() for DEGs in Mic.3 were diagramed in the KEGG pathway ‘Cytokine-cytokine receptor interaction’ and ‘Cholesterol metabolism’ (Supplemental Figure 6).

### Compositional analysis of bulk RNA-seq

Digital deconvolution was performed using ROSMAP, Mayo, and the Knight ADRC datasets was previously described^72,79^. Briefly, gene markers were selected for major brain cell-types: neurons, astrocytes, microglia, and oligodendrocytes. The *CellMix*^80^ was used to estimate the relative proportion of four cell types in each sample.

AD and controls in ROSMAP, Mayo, and Knight ADRC were selected for this analysis. Subjects with outliners (defined by Tukey’s fences) in any of the four cell types were removed from each study before analysis. The effect of risk and protective variants on cell-type proportion was calculated by the generalized linear model (GLM) *glm*(). The model also listed age, sex, AD status, PC1 and PC2 derived from GWAS data, TIN, PMI, and the remaining risk/protective variant as covariates. A threshold was set as *p*-value < 0.05. The following model was used:

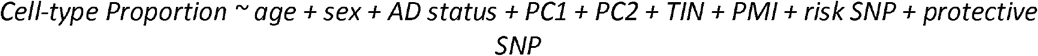

### Compositional analysis of single-nucleus RNA-seq

Microglia proportion was calculated relative to total cell count in each donor. The effect of risk and protective variants on microglia proportion was evaluated by *GLM glm*(). The model listed age, sex, AD status, PC1 and PC2 derived from GWAS data, PMI and the alternative risk/protective variant as covariates. A threshold was set as *p*-value < 0.05. The following model was used:

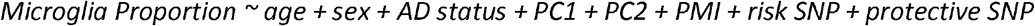

Microglia cluster proportion was calculated relative to total microglia cell count in each donor. The effect of risk and protective variant on microglia cluster proportion was evaluated by GLM *glm*(). The model listed age, sex, AD status, PC1 and PC2 derived from GWAS data, PMI, and the remaining risk/protective variant as covariates. A threshold was set as *p*-value < 0.05. The following model was used:

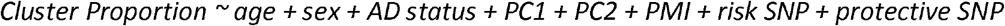

To verify that the increment of Mic.3 in risk variant is the driving force for overall microglia increase. Microglia proportion amongst all cell types was recalculated in GLM *glm*() after dropping Mic.3 from all samples (Supplemental Figure 7).

### Integration analysis with induced transcription factor microglia-like cells (iTF-Microglia)

The single cell RNA-seq dataset for iTF-microglia was downloaded from a publicly available database^35^. To integrate the human microglia with iTF-microglia, ADAD samples were removed from the human dataset. Both datasets were normalized using *SCTransform* to regress out the variance caused by the total number of the molecules detected (nCount_RNA) and the gene detected (nFeature_RNA) in each cell.

Dataset integration was performed following Seurat’s best practices workflow^81^. Human microglia from the Knight ADRC snRNA-seq were set as the reference by *FindIntegrationAnchors* and *IntegrateData*. Integration features were set at 3000 using *SelectIntegrationFeatures* and *PrepSCTintegration*. The Mutual Nearest Neighbours (MNN) RunFastMNN ^82^ was used as a graphic reduction method. The UMAP and integrated clusters are defined by *FindNeighbors* (PC = 1:20) and *FindClusters* (resolution = 0.2). The integrated map showed the relative location overlap between human Knight ADRC microglia and iTF-microglia. The top 50 DEGs for Microglia cluster 3 (Mic.3) were selected and visualized to display the gene expression pattern between clusters in integrated human brain microglia and iTF-microglia (Supplemental Figure 8).

### Cell culture and quantitative assays

#### Primary mouse microglia culture

Primary mouse microglia cultures were obtained from mixed cultures prepared from the hippocampi and cerebral cortices of WT and *Ch25h* KO mice on postnatal day (P) 1-3. Microglial cells were isolated by shaking flasks for 45 min at 230 rpm on day ten after plating. Cells were then seeded on poly-L-ornithine (Sigma) pre-coated wells in DMEM containing 20% heat-inactivated fetal bovine serum (FBS).

### Human macrophage culture and lentivirus transduction

The culture methods for human macrophage and lentivirus vectors used in this experiment were previously described^10^. Briefly, peripheral blood mononuclear cells (PBMCs) were purified from human blood on Ficoll-Paque PLUS density gradient (Amersham Biosciences, Piscataway, NJ). PBMCs were cultured in six-well culture plates (3 × 10^6^ cells per well) in RPMI 1640 without FBS to generate macrophages. After 2 hours of culture, PBMCs were washed twice with 1X PBS and cultured in RPMI supplemented with macrophage colony-stimulating factor (MCSF) (50 ng/ml) for seven days at 37°C with 5% CO2. Supernatants from macrophage cultures were collected, centrifuged at 21,000g for 15 min, and filtered through a 0.22-μm filter to remove all cells and membrane debris. Macrophage supernatants were frozen in aliquots at −80°C until used in the ELISA for human sTREM2. Lentiviral vectors encoding GFP (PS100071) and human *MS4A4A* WT (NM_148975) in Myc-DDK-tagged vector (RC204646L1) were obtained from OriGene. All constructs were verified by Sanger sequencing. The viral particles were transfected and harvested from the human embryonic kidney (HEK) 293T cells. Cultured monocyte-derived macrophages were transduced with lentivirus vector for 24 hours and then rinsed, and fresh media was added. Cells and media were collected 48 hours later for sTREM2 ELISA and quantitative polymerase chain reaction (qPCR) analyses.

### Mouse sTrem2 measurement

Mouse sTrem2 was measured as following. Briefly, an anti-mouse Trem2 antibody (MAB1729, R&D Systems, [2mg/ml]) was used as a capture antibody and coated overnight at 4°C on MaxiSorp 96-well plates (Nalge Nunc International, Rochester, NY) in sodium bicarbonate coating buffer [0.015 M Na2CO3and 0.035 M NaHCO3 (pH 9.6)]. All the washes between the different steps were done four times with PBS/0.05% Tween 20 (Sigma-Aldrich). After washing, wells were blocked for 4 hours at room temperature (RT) with phosphate-buffered saline (PBS)*/*3% BSA. Freshly thawed supernatant and recombinant mouse Trem2 standards (msTrem2-Fc (R&D Systems #1729-T2) were incubated in duplicate overnight at 4°C. For detection, wells were incubated with a Trem2 biotinylated detection antibody (R&D Systems, BAF1729, [0.25μg/ml]) for 1 hour at RT on an orbital shaker. After washing, wells were incubated with horseradish peroxidase-labeled streptavidin (BD Biosciences, San Jose, CA) for 1 hour at RT with orbital shaking. Horseradish peroxidase visualization was performed with 3,3’,5,5’ tetramethyl-benzidine (Sigma-Aldrich, St. Louis, MO) added to each well for 10 min at RT in the dark. Color development was stopped by adding an equal volume of 2.5N H_2_SO_4_. Optical density of each well was determined at 450 nm.

### Quantitative PCR

The effect of overexpression was measured by real-time qPCR analysis. Macrophages transduced with the lentiviral particles were collected in TRIzol reagent (Thermo Fisher Scientific), and total RNA was extracted using the RNeasy Mini Kit (Qiagen). cDNA was prepared from the total RNA using the High-Capacity cDNA Reverse Transcription Kit (Thermo Fisher Scientific). Gene expression levels were analyzed by real-time PCR using TaqMan assays for *MS4A4A* (Hs00254780_m1), *TREM2* (Hs00219132_m1), *CH25H* (Hs02379634_s1), *ATP6V0D2* (Hs00403032_m1), *LAMP2* (Hs00174474_m1), *CASP1* (Hs00354836_m1), *NLRP1* (Hs00248187_m1), *CCL2* (Hs00234140_m1), and *GAPDH* (Hs02758991_g1) on a QuantStudio 12K Flex Real-Time PCR System (Thermo Fisher Scientific). To avoid amplification interference, expression assays were run in separate wells from the housekeeping gene GAPDH. Real-time data were analyzed by the comparative C_T_ method. Average C_T_ values for each sample were normalized to the average C_T_ values for the housekeeping gene *GAPDH*.

### Calcium Assay

THP-1 monocytes were cultured and passaged in T75 flask and differentiated into macrophages using two doses of 20ng/mL PMA treatment every two days. On the fourth day, mature THP-1 macrophages were harvested and seeded into a 96-well plate at a density of 50K cells/well. After allowing cells to adhere, cells were treated with 2 pmol/well of scrambled (ThermoFisher Cat. No. 4390843) or *MS4A4A* silencer s27981 (ThermoFisher Cat. No. 4392420) were treated following standard protocol provided by Lipofectamine RNAiMAX (Thermo Fisher Cat. No. 13778150). Media was removed and washed with 1X PBS three days after treatment. The macrophages were incubated at 37°C with 50μL Buffer + 50μL Fluo-4 (Fluo-4 Direct™ Calcium Assay Kit Cat. No. F10472) and incubated for 1 hour. After incubation, the baselines were read with a BioTek Synergy plate reader at 494 nm Excitation/516nm Emission. The kinetics of calcium stimulants were recorded immediately after adding SDF1α (1μg/mL; PeproTech Cat. No. AF-300-28A) or ionomycin (1μM; STEMCELL Technologies Cat. No. 73722) for 300 seconds. Analyses were performed with the proprietary software Gen5.

## Data Availability

All data used in this study is from publicly available sources. All data produced in the present study are available upon reasonable request to the authors.

## Acknowledgements

We would like to thank the research participants and their families who generously participated in this study. We thank Torri Ball and Grant Galasso for thoughtful discussions. This work was supported by access to equipment made possible by the Hope Center for Neurological Disorders, the Neurogenomics and Informatics Center, and the Departments of Neurology and Psychiatry at Washington University School of Medicine. Funding provided by the National Institutes of Health (AG067764, AG005681, AG062734, AG032438, AG058501, AG071706, NS118146 and NS127211), Hope Center for Neurological Disorders, Chan Zuckerberg Initiative (CMK, OH, MK), Thome Memorial Foundation, BrightFocus Foundation, and UL1TR002345, Alzheimer’s Association ZEN-22-969903 (MK). The recruitment and clinical characterization of Knight ADRC research participants at Washington University were supported by NIH P30AG066444 (JCM), P01AG03991 (JCM), and P01AG026276 (JCM). Diagrams were generated using BioRender.com.

Study data were provided by the Rush Alzheimer’s Disease Center, Rush University Medical Center, Chicago. Data collection was supported through funding by NIA grants P30AG10161 (ROS), R01AG15819 (ROSMAP; genomics and RNAseq), R01AG17917 (MAP), R01AG30146, R01AG36042 (5hC methylation, ATACseq), RC2AG036547 (H3K9Ac), R01AG36836 (RNAseq), R01AG48015 (monocyte RNAseq) RF1AG57473 (single nucleus RNAseq), U01AG32984 (genomic and whole exome sequencing), U01AG46152 (ROSMAP AMP-AD, targeted proteomics), U01AG46161(TMT proteomics), U01AG61356 (whole genome sequencing, targeted proteomics, ROSMAP AMP-AD), the Illinois Department of Public Health (ROSMAP), and the Translational Genomics Research Institute (genomic). Additional phenotypic data can be requested at www.radc.rush.edu. Genotype data: doi:10.1038/mp.2017.20. RNAseq: doi:10.1038/s41593-018-0154-9.

Data collection and sharing for this project was supported by The Dominantly Inherited Alzheimer Network (DIAN, U19AG032438) funded by the National Institute on Aging (NIA),the Alzheimer’s Association (SG-20-690363-DIAN), the <u>German Center for Neurodegenerative Diseases</u> (DZNE), Raul Carrea Institute for Neurological Research (FLENI), Partial support by the Research and Development Grants for Dementia from Japan Agency for Medical Research and Development, AMED, and the Korea Health Technology R&D Project through the Korea Health Industry Development Institute (KHIDI), Spanish Institute of Health Carlos III (ISCIII), Canadian Institutes of Health Research (CIHR), Canadian Consortium of Neurodegeneration and Aging, Brain Canada Foundation, and Fonds de Recherche du Québec – Santé. This manuscript has been reviewed by DIAN Study investigators for scientific content and consistency of data interpretation with previous DIAN Study publications. We acknowledge the altruism of the participants and their families and contributions of the DIAN research and support staff at each of the participating sites for their contributions to this study.

