## Supplemental Figures for "MS4A4A modifies the risk of Alzheimer disease by regulating lipid metabolism and immune response in a unique microglia state"

### Supplemental Figure 1

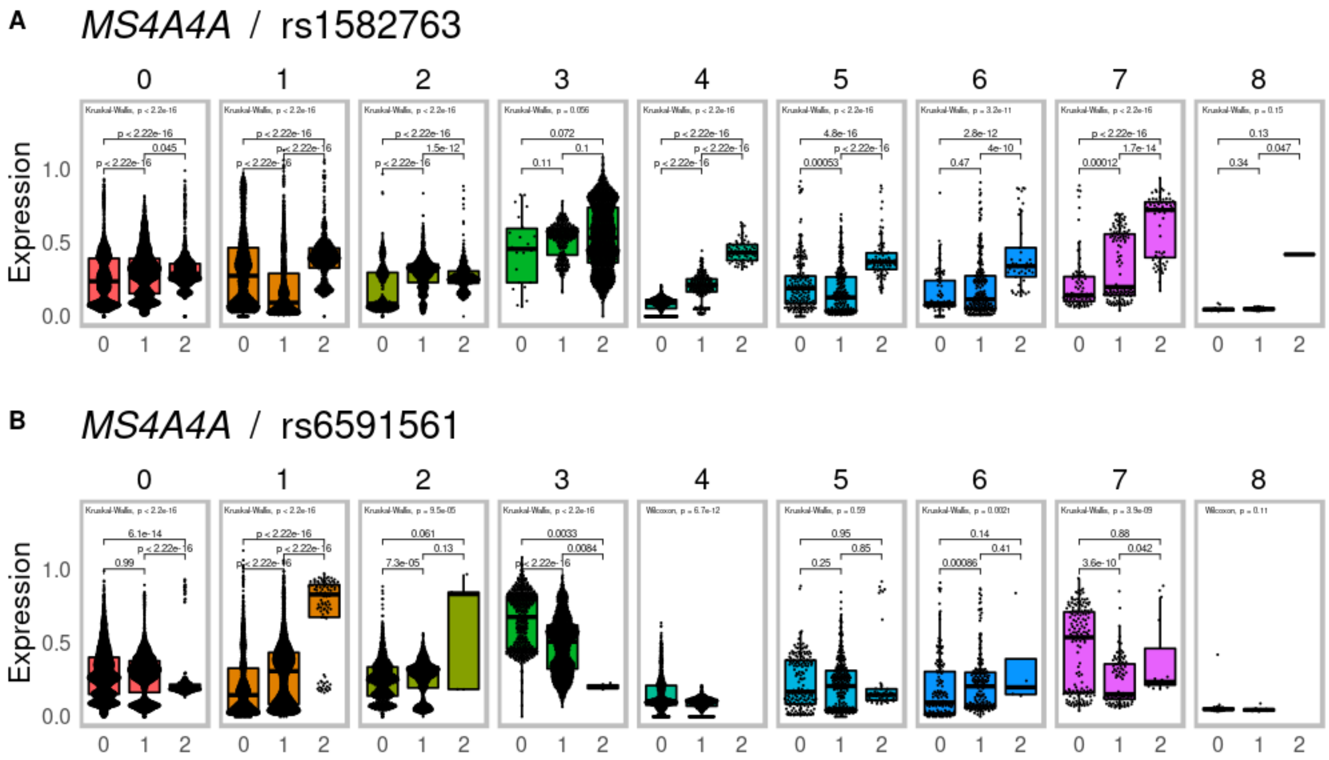


**Supplemental Figure 1. MS4A4A expression split by (A) rs1582763 and (B) rs6591561 genotypes across microglia clusters Mic.0, Mic.1, Mic.2.**

### Supplemental Figure 2

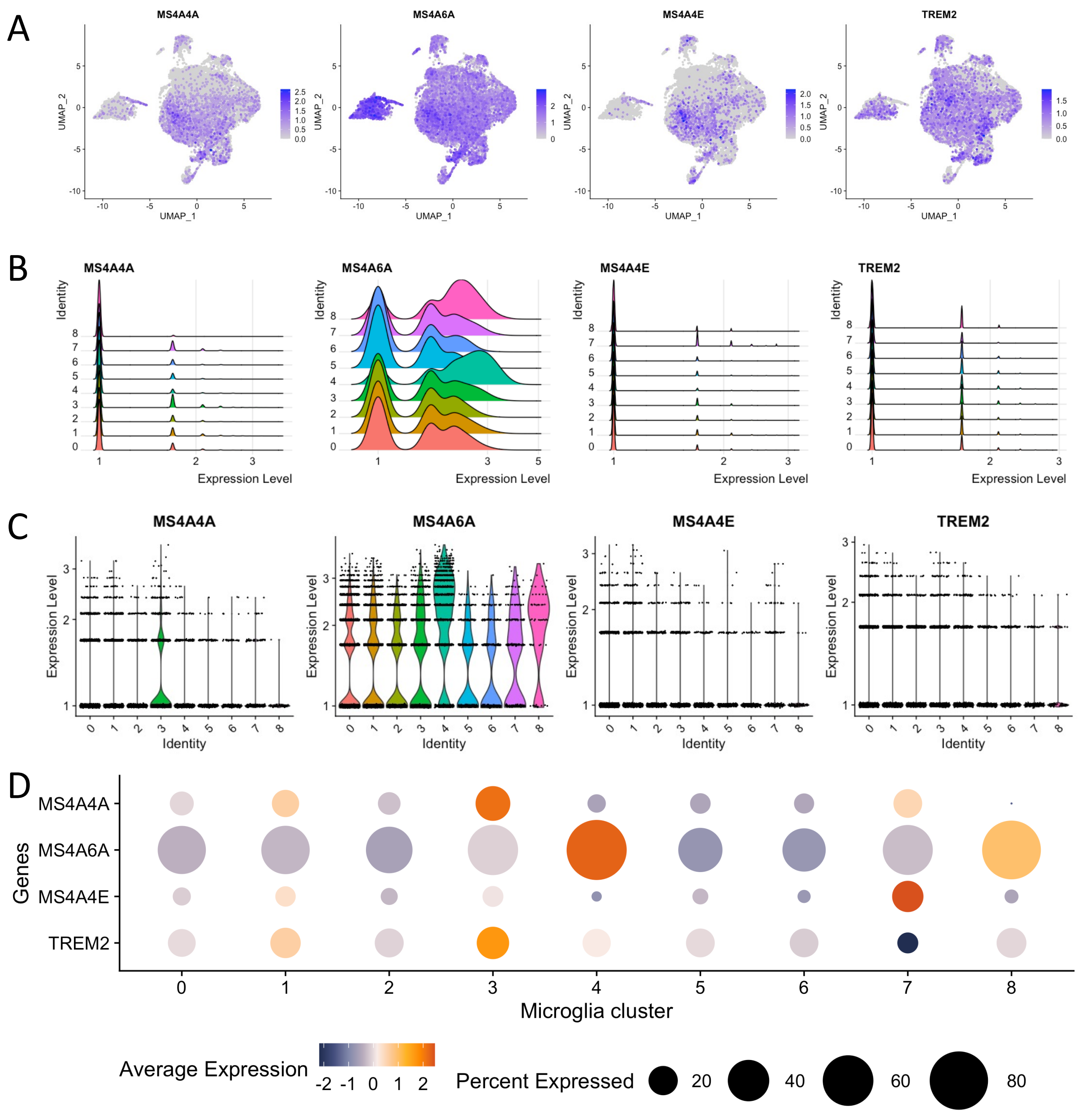


**Supplemental Figure 2. *MS4A4A*, *MS4A6A*, *MS4A4E* and *TREM2* expression across the microglia clusters in human brain snRNA-seq.** (A) Feature plot (B) Ridge plot (C) Violin plot (D) Dot plot of *MS4A4A*, *MS4A6A*, *MS4A4E* and *TREM2* across different microglia clusters.

### Supplemental Figure 3

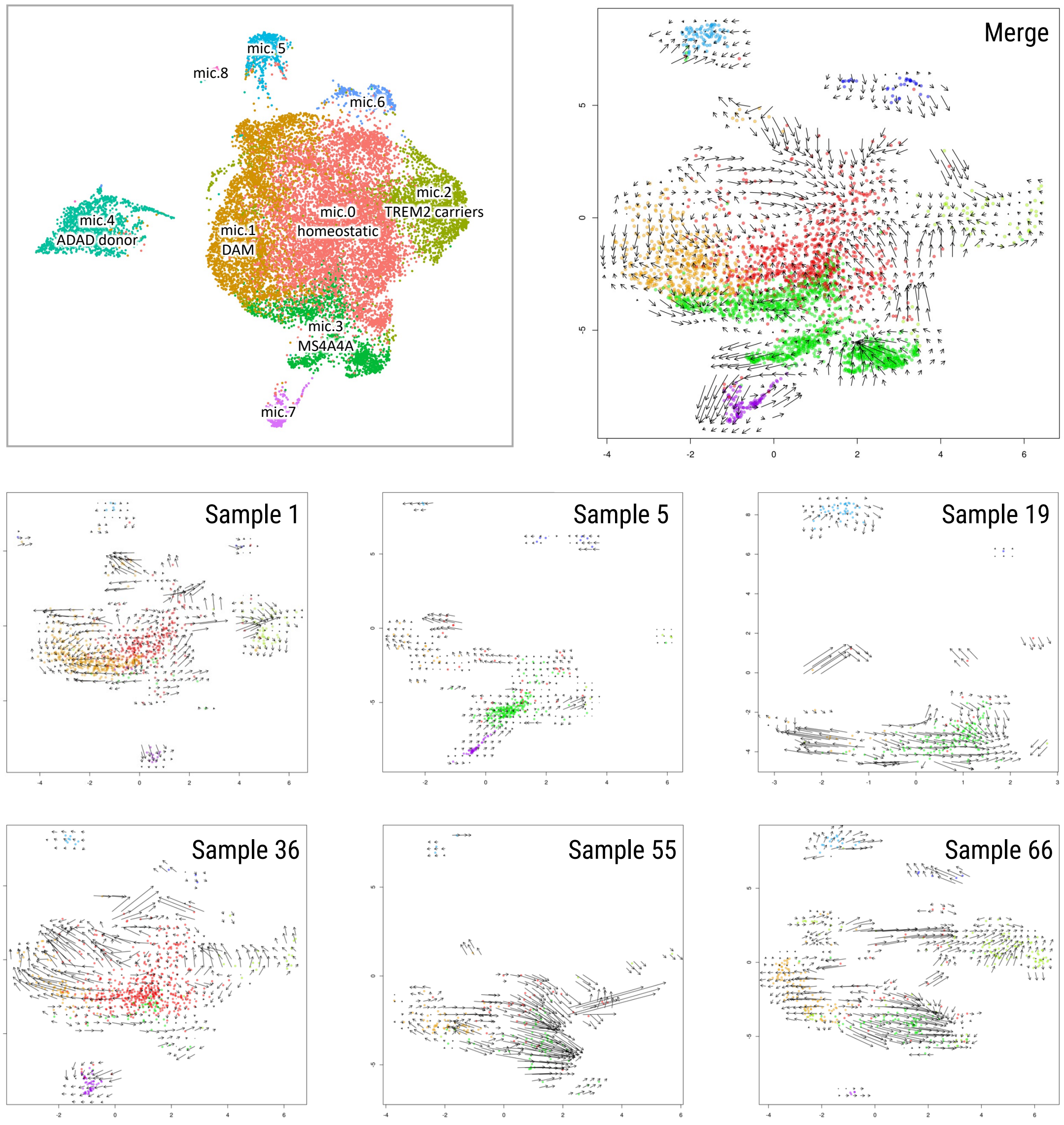


**Supplemental Figure 3. Examples of RNA velocity in some individual samples that contains Mic3.** Top row represents merged data. Individual donors represents below.

### Supplemental Figure 4

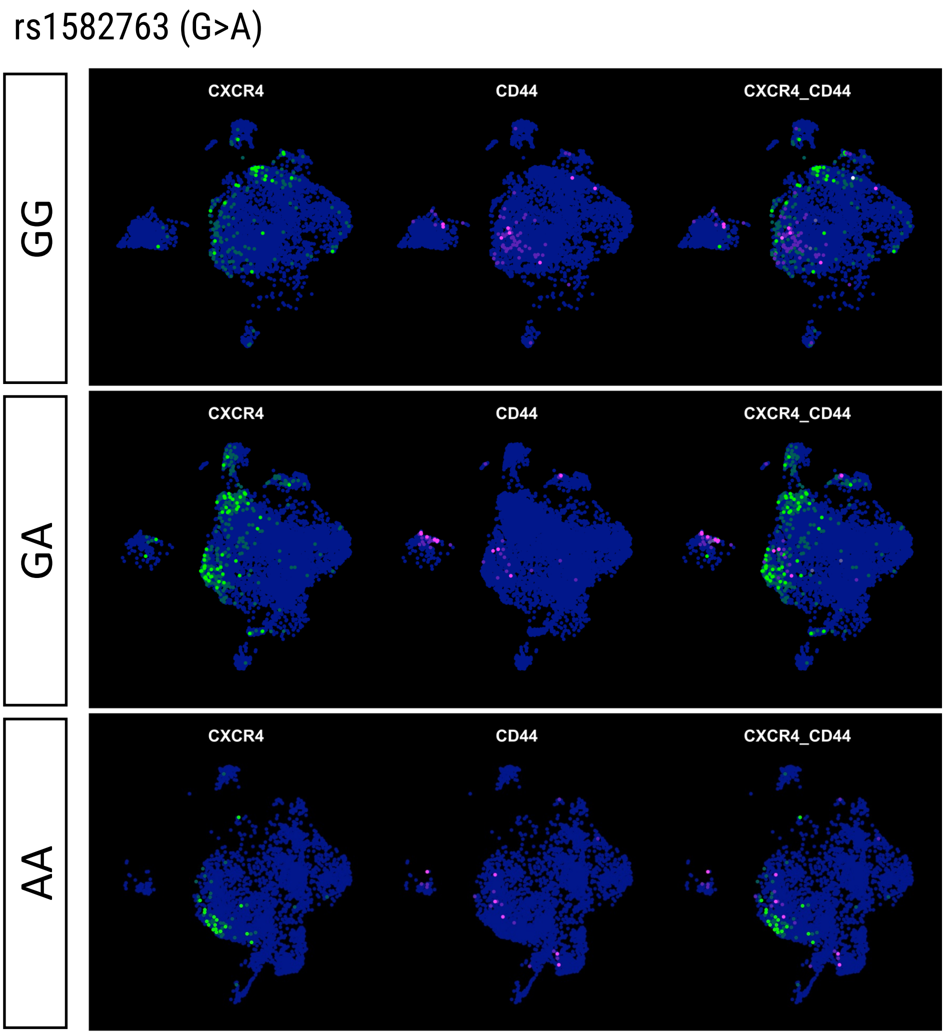


**Supplemental Figure 4. Low CXCR4 and CD44 co-expression in human brain snRNAseq data.** rs1582763-A minor allele carriers have less CD44+ nuclei and more CXCR4+ nuclei in microglia.

### Supplemental Figure 5

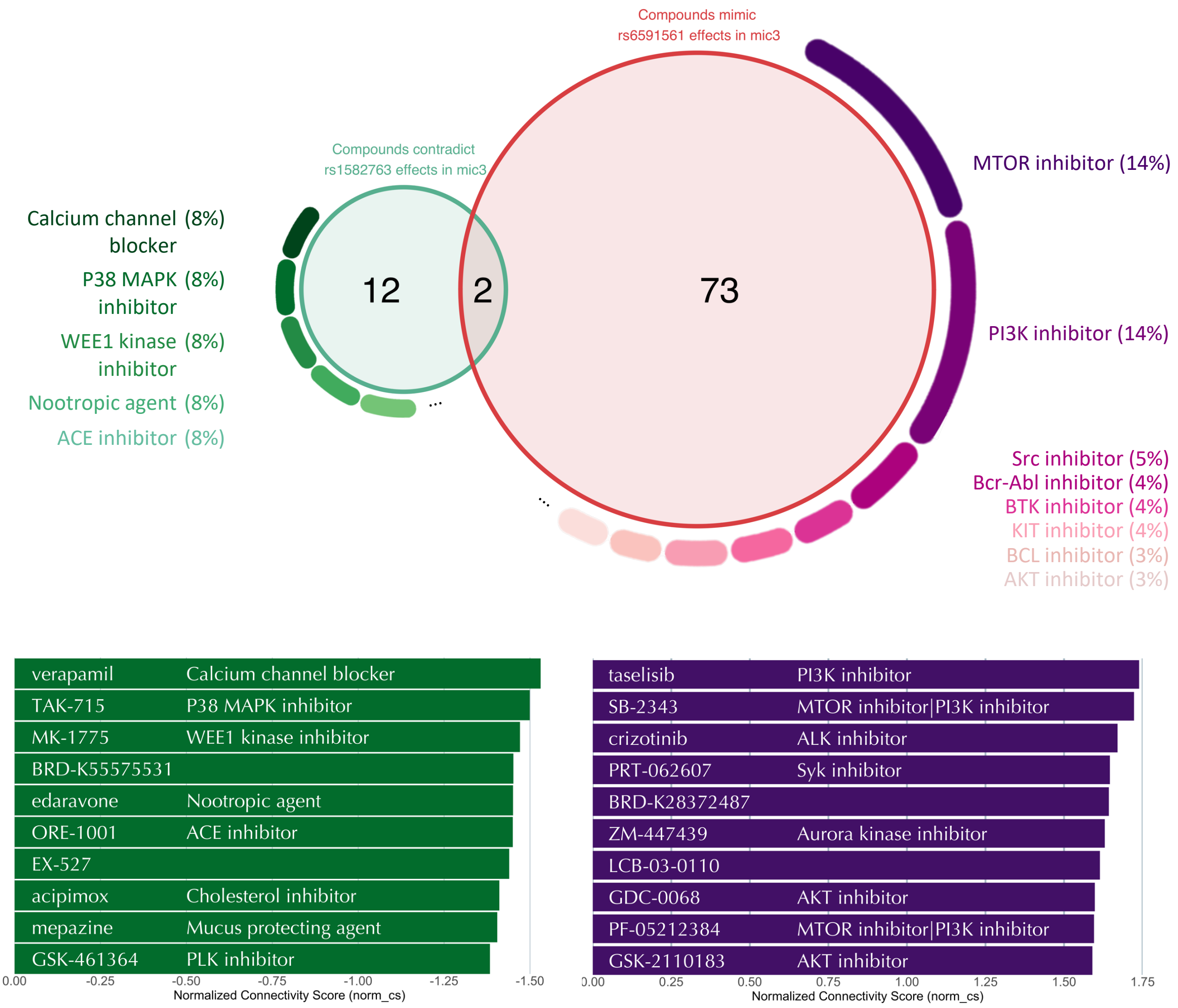


**Supplemental Figure 5. CMAP predicted compounds which contradict 1582763 and mimic rs6591561 in Mic3**. (Left) Compounds with effects that reverse gene expression changes associated with the protective variant rs1582763. (Right) Compounds with effects that mimic gene expression changes associated with the risk variant rs6591561.

### Supplemental Figure 6

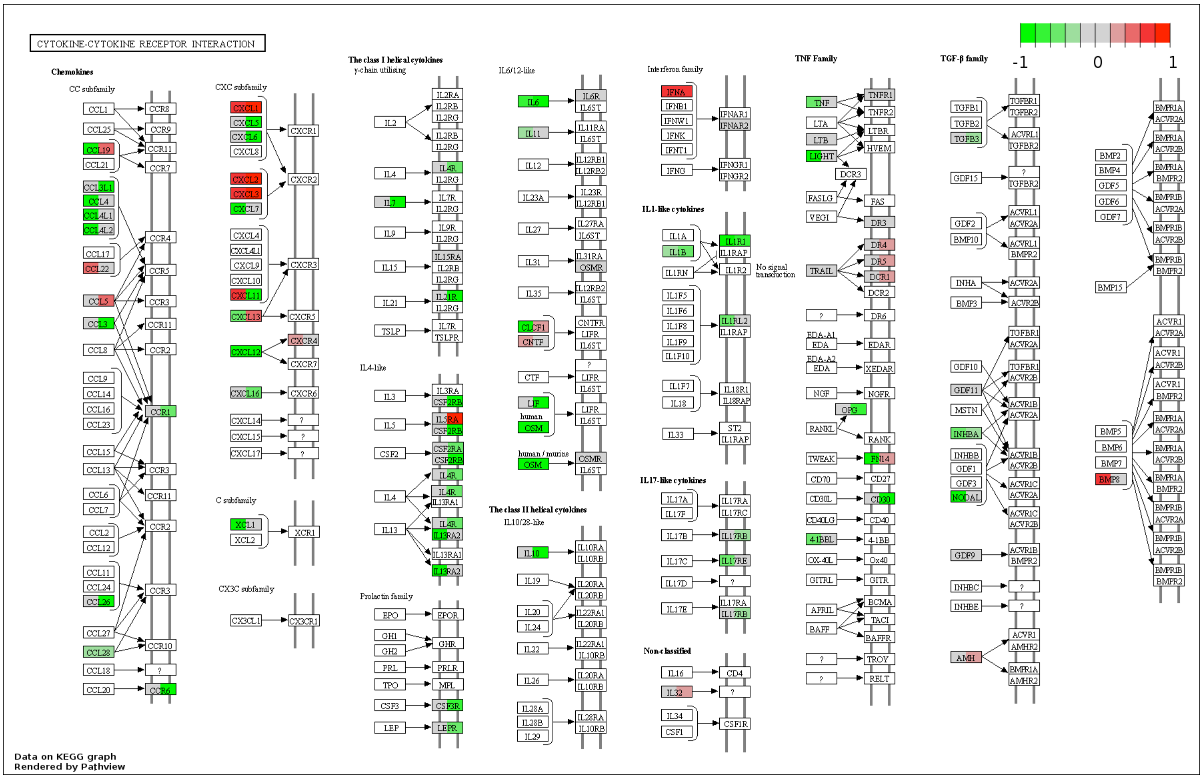

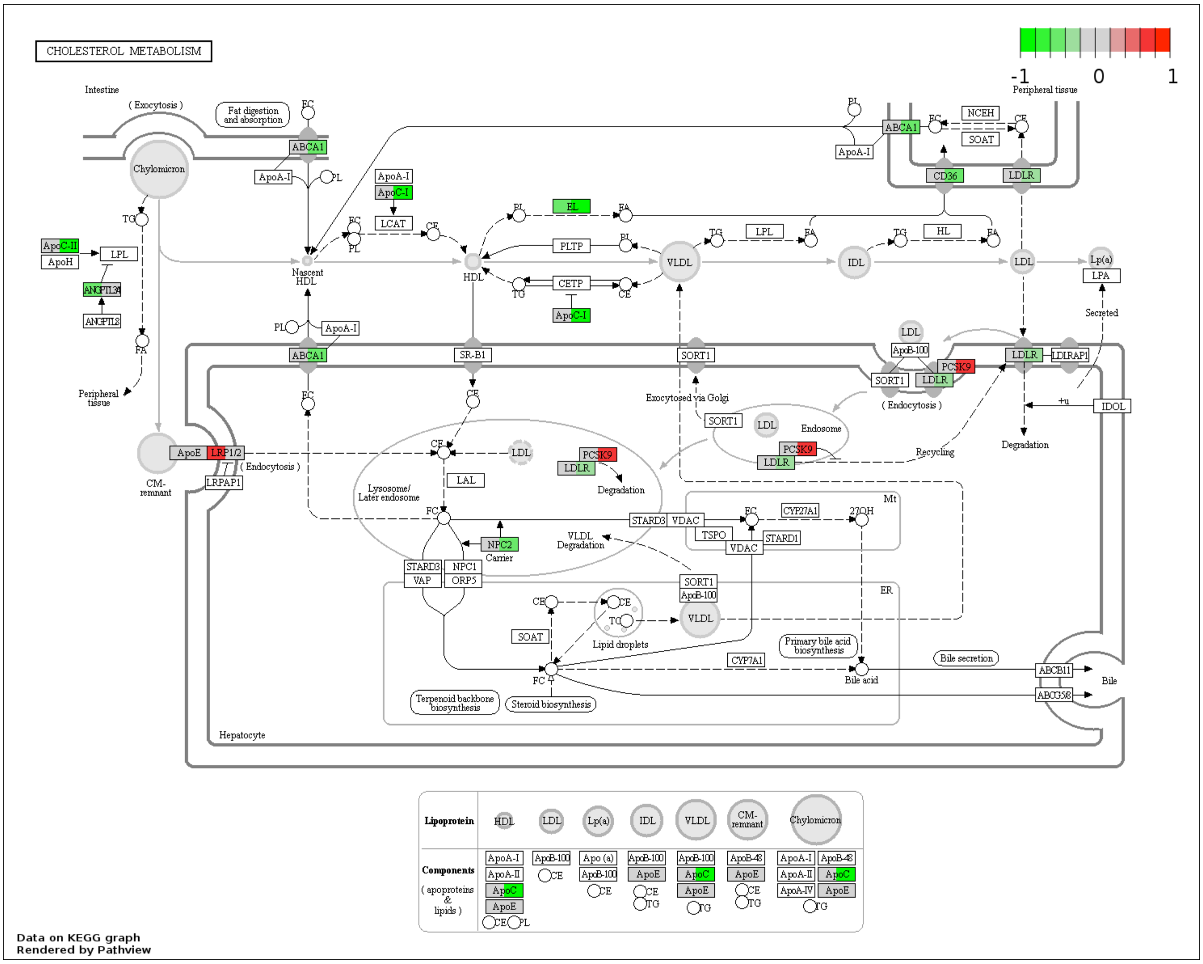

**Supplemental Figure 6**. **Pathways for “Cytokine-cytokine receptor interaction” and “Cholesterol metabolism” for differentially expressed genes in meta-analysis of bulk RNA-seq data in human brains.** For each given box, left is rs1582763, right is rs6591561. Red and green indicate up-regulated and down-regulated DEG in LogFC.

### Supplemental Figure 7

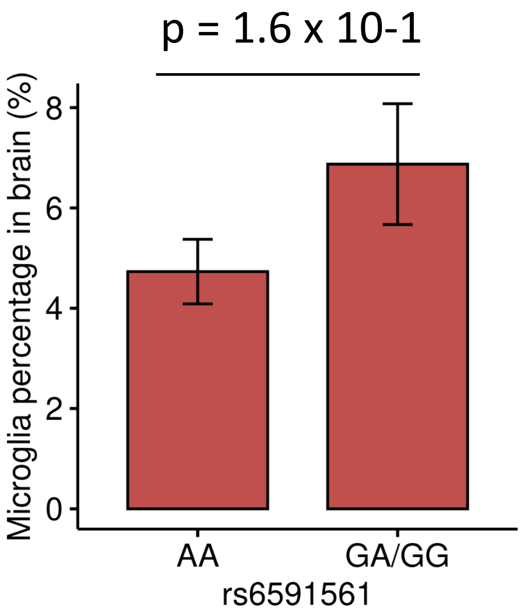


**Supplemental Figure 7. Removing Mic.3 eliminated the effect of rs6591561 on overall microglia population.**

### Supplemental Figure 8

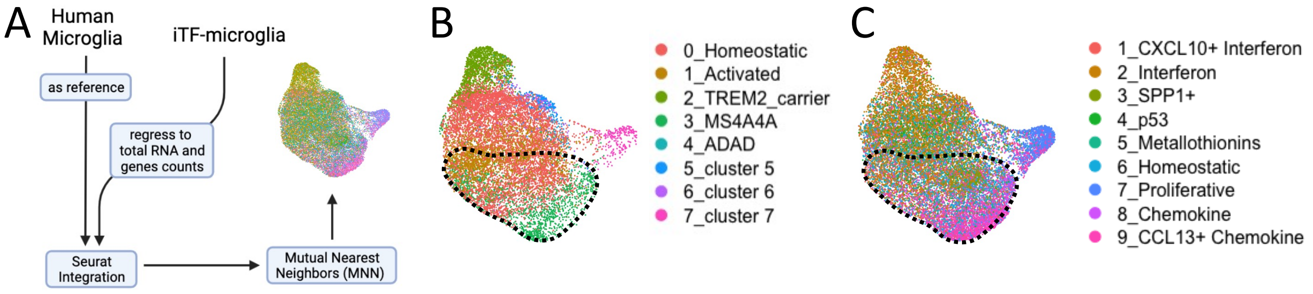


**Supplemental Figure 8**. **Integration of human microglia and iTF-microglia data.** A. Schematic diagram of transcriptomic integration with human microglia and iTF-microglia using the Mutual Nearest Neighbors (MNN) method. (B) UMAP of Knight ADRC clusters. (C) iTF-microglia cluster clusters. Circle shows overlapping area between Mic.3 and iTF-microglia.8 (Chemokine) and iTF-microglia.9 (CCL13+ Chemokine).
